# AZD7442 demonstrates prophylactic and therapeutic efficacy in non-human primates and extended half-life in humans

**DOI:** 10.1101/2021.08.30.21262666

**Authors:** Yueh-Ming Loo, Patrick M. McTamney, Rosalinda H. Arends, Robert A. Gasser, Michael E. Abram, Anastasia Aksyuk, Seme Diallo, Daniel J. Flores, Elizabeth J. Kelly, Kuishu Ren, Richard Roque, Kim Rosenthal, Katie Streicher, Kevin M. Tuffy, Nicholas J. Bond, Owen Cornwell, Jerome Bouquet, Lily I. Cheng, James Dunyak, Yue Huang, Anton I. Rosenbaum, Hanne Andersen, Robert H. Carnahan, James E. Crowe, Ana I. Kuehne, Andrew S. Herbert, John M. Dye, Helen Bright, Nicole L. Kallewaard, Menelas N. Pangalos, Mark T. Esser

**Affiliations:** Virology and Vaccine Discovery, Microbial Sciences, BioPharmaceuticals R&D, AstraZeneca, Gaithersburg, MD, USA; Clinical Pharmacology and Quantitative Pharmacology, Microbial Sciences, Clinical Pharmacology and Safety Sciences, BioPharmaceuticals R&D, AstraZeneca, Gaithersburg, MD, USA; Clinical Development, Microbial Sciences, BioPharmaceuticals R&D, AstraZeneca, Gaithersburg, MD, USA; Translational Medicine, Microbial Sciences, BioPharmaceuticals R&D, AstraZeneca, Gaithersburg, MD, USA; Analytical Sciences, BioPharmaceuticals R&D, AstraZeneca, Granta Park, Cambridge, UK; Integrated Bioanalysis, Clinical Pharmacology and Quantitative Pharmacology, Clinical Pharmacology and Safety Sciences, R&D, AstraZeneca, San Francisco, CA, USA; Oncology Safety Pathology, Clinical Pharmacology and Safety Sciences, BioPharmaceuticals R&D, AstraZeneca, Gaithersburg, MD, USA; Clinical Pharmacology and Pharmacometrics, Microbial Sciences, BioPharmaceuticals R&D, AstraZeneca, Gaithersburg, MD, USA; BIOQUAL Inc., Rockville, MD, USA; Department of Pediatrics, Vanderbilt University Medical Center, Nashville, TN, USA; The Vanderbilt Vaccine Center, Vanderbilt University Medical Center, Nashville, TN, USA; Department of Pathology, Microbiology and Immunology, Vanderbilt University, Nashville, TN, USA; USAMRIID, Fort Detrick, MD, USA; BioPharmaceuticals R&D, AstraZeneca, Cambridge, UK; Microbial Sciences, BioPharmaceuticals R&D, AstraZeneca, Gaithersburg, MD, USA

## Abstract

Despite the success of SARS-CoV-2 vaccines, there remains a need for more prevention and treatment options for individuals remaining at risk of COVID-19. Monoclonal antibodies (mAbs) against the viral spike protein have potential to both prevent and treat COVID-19, and reduce the risk of severe disease and death. Here, we describe AZD7442, a combination of two mAbs, AZD8895 (tixagevimab) and AZD1061 (cilgavimab), that simultaneously bind to distinct non-overlapping epitopes on the spike protein receptor binding domain to potently neutralize SARS-CoV-2. Initially isolated from individuals with prior SARS-CoV-2 infection, the two mAbs were designed to extend their half-lives and abrogate effector functions. The AZD7442 mAbs individually prevent the spike protein from binding to angiotensin-converting enzyme 2 receptor, blocking virus cell entry. Together, these two mAbs create a higher barrier to viral escape and a wider breadth of coverage, neutralizing all known SARS-CoV-2 variants of concern. In a non-human primate model of SARS-CoV-2 infection, prophylactic AZD7442 administration prevented infection, while therapeutic administration accelerated virus clearance from lung. In an ongoing Phase I study in healthy participants (NCT04507256), 300 mg intramuscular AZD7442 provided SARS-CoV-2 serum geometric mean neutralizing titers >10-fold above those of convalescent sera for ≥3 months, which remained 3-fold above those of convalescent sera 9 months post-AZD7442 administration. Approximately 1–2% of serum AZD7442 levels were detected in nasal mucosa, a site of SARS-CoV-2 infection. Extrapolation of the time course of serum AZD7442 concentrations suggests AZD7442 may provide up to 12 months of protection and benefit individuals at high-risk of COVID-19.

## Introduction

The coronavirus disease 2019 (COVID-19) pandemic caused by severe acute respiratory syndrome coronavirus 2 (SARS-CoV-2) continues to cause substantial morbidity and mortality world-wide. While roll out of effective COVID-19 vaccines has reduced hospitalizations and death in several countries (*1, 2*), SARS-CoV-2 infection continues to spread globally, as variants with increased transmissability and immune evasion continue to emerge. The need for new therapies to protect individuals who remain at risk of COVID-19 persists, which includes unvaccinated individuals, individuals who are unable to mount an adequate immune response following vaccination (*3–7*), and individuals with breakthrough infections despite full vaccination (*8*).

SARS-CoV-2-neutralizing monoclonal antibodies (mAbs) represent an approach for both the prevention and/or treatment of COVID-19 (*9, 10*). The receptor binding domain (RBD) of the spike protein of SARS-CoV-2 mediates attachment to human angiotensin-converting enzyme 2 (ACE2) receptor resulting in viral entry into host cells (*10*). Many individuals with SARS-CoV-2 infection develop neutralizing antibodies to the spike protein (*11, 12*), which correlate with protection against symptomatic infection (*13*). Antibodies targeting the spike protein have also been shown to limit the progression of SARS-CoV-2 infection and the development of COVID-19 (*14–16*).

AZD7442 is a combination of two fully human, long-acting SARS-CoV-2-neutralizing antibodies, AZD8895 (tixagevimab) and AZD1061 (cilgavimab), in clinical development for the prevention of symptomatic COVID-19 and the treatment of mild-to-moderate and severe COVID-19 (*17, 18*). AZD8895 and AZD1061 were derived from the B cells of individuals with prior SARS-CoV-2 infection (*19*). Their progenitor mAbs were shown to potently and synergistically neutralize SARS-CoV-2 *in vitro* and confer protection in animal models of SARS-CoV-2 infection when co-administered (*19*), supporting further development of this combination. The variable regions of the progenitor mAbs were reformatted as immunoglobulin 1 kappa (IgG1κ) with amino acid substitutions in the fragment crystallizable (Fc) regions to extend their serum half-lives (*20, 21*) and reduce Fc gamma receptor (FcγR) and complement binding, to create AZD8895 and AZD0161 (*22*).

In this study we describe the preclinical and translational characteristics of AZD8895, AZD1061, and AZD7442, and evaluate the potential of AZD7442 to both prevent and treat SARS-CoV-2 infection in non-human primate (NHP) models. Additionally, we characterize AZD7442 pharmacokinetics and transudation to the nasal mucosae in healthy adult participants enrolled in a Phase 1 clinical study .

## Results

### AZD7442 antibodies AZD8895 and AZD1061 simultaneously bind the SARS-CoV-2 spike protein with high affinity

Co-crystal structures of AZD8895 and AZD1061 showed simultaneous binding to the RBD at distinct, non-contiguous and non-overlapping epitopes (**Fig. 1A**). Hydrogen-deuterium exchange mass spectrometry confirmed that AZD8895 and AZD1061 associate with distinct peptide residues on opposing faces of the RBD **(Fig. S1)**. AZD8895, AZD1061 and AZD7442 bound to the SARS-CoV-2 spike protein with high affinity (*K*_D_ 2.8, 13.0 and 13.7 pM, respectively). Compared with hACE2, AZD7442 had a >3,000-fold higher affinity for the spike protein (*K*_D_ 43,000 pM) (**Fig. 1B**). AZD8895 and AZD1061 potently blocked RBD binding to ACE2 individually (IC_50_ 47.7 and 79.6 ng/mL, respectively) (**Fig. 1C**), demonstrating that the two antibodies can independently block RBD binding to ACE2.

**Figure 1.**
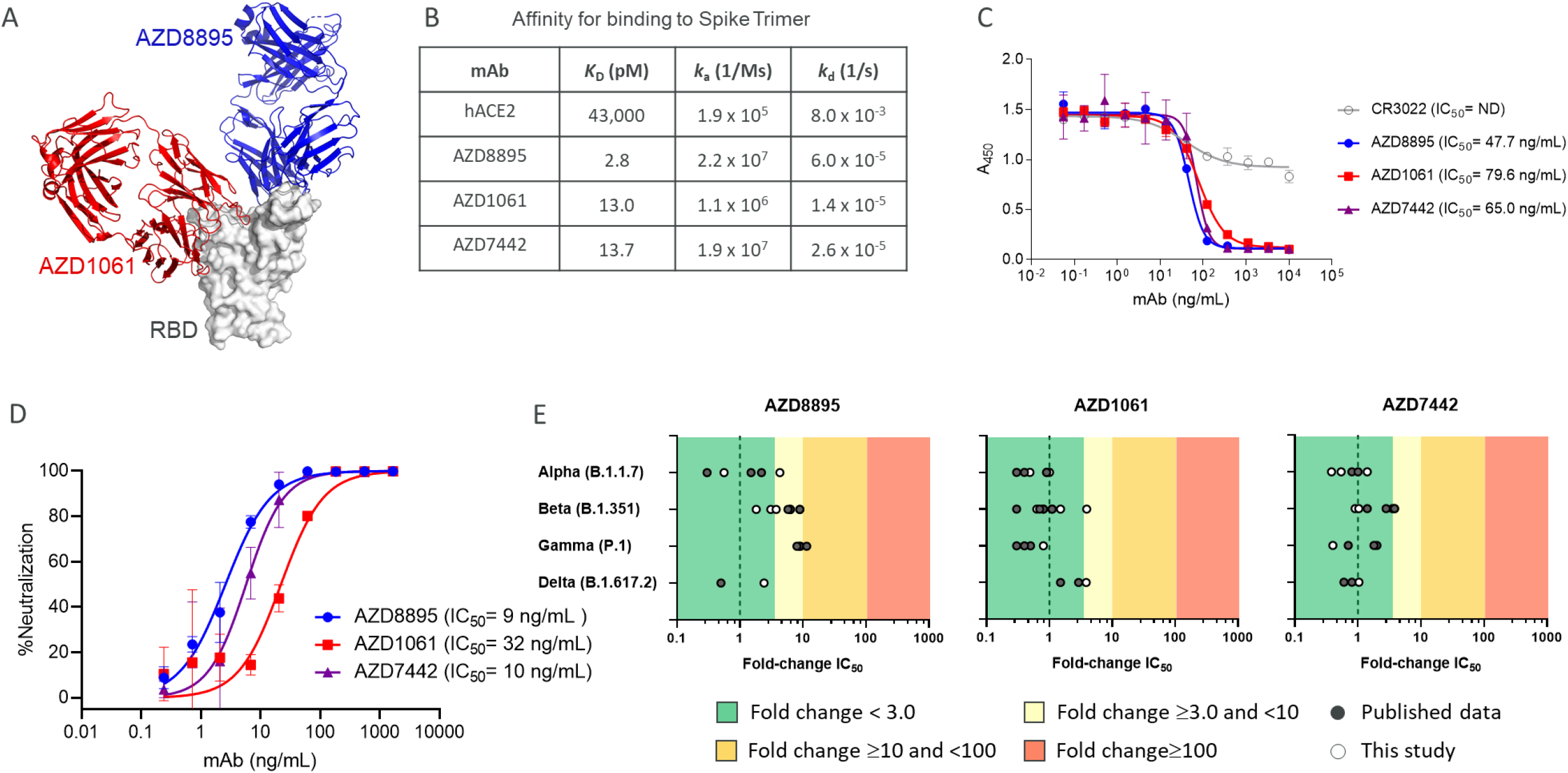
Simultaneous binding of AZD1061 and AZD8895 (AD7442) to the SARS-CoV-2 RBD blocks binding to ACE2 and potently neutralizes SARS-CoV-2 variants of concern. **A.** AZD8895 and AZD1061 simultaneously bind to distinct, non-overlapping epitopes on the spike protein RBD and sterically block RBD binding to ACE2. Side-view depiction showing cartoon representations of AZD8895 (blue) and AZD1061 (red) on top of the RBD (white) in surface representation based on co-crystal structures of AZD8895 and AZD1061 with RBD (*24*). **B.** AZD8895, AZD1061, and AZD7442 binding kinetics on the SARS-CoV-2 S trimer (S2P ectodomain protein). Data shown are from a representative experiment from at least two independent assays (*48*). **C.** AZD8895, AZD1061, and AZD7442 block binding of the RBD to ACE2. Measurements were taken across a series of mAb concentrations and the resulting non-linear regression curves were used to calculate IC_50_ values. Data shown were performed in duplicate and represent at least two independent assays. **D.** AZD8895, AZD1061, and AZD7442 neutralize the USA-WA1/2020 strain of SARS-CoV-2 *in vitro* . Non-linear regression dose-response curves from a representative experiment are shown, with mean and SD error from two or more technical replicates. Mean IC_50_ values were calculated from three independent experiments. **E.** AZD8895, AZD1061, and AZD7442 neutralize SARS-CoV-2 VOC *in vitro*. Data shown represent fold-change in neutralization potencies (IC_50_) of AZD8895, AZD1061 and AZD7442 against the Alpha (B.1.1.7), Beta (B.1.351), Gamma (P.1), and Delta (B.1.617.2) VOC compared to the USA-WA1/2020 or AUS/VIC01/2020 reference strains. Data shown in solid circles have been published previously (*24, 28, 29, 31, 33-35*). Data shown in open circles is from this study, performed at PHE (Alpha, Beta, Gamma, and Delta), USAMRIID (Alpha and Beta), and IRF/NIAID (Alpha and Beta). ACE2, angiotensin-converting enzyme 2; IC_50_, half maximal inhibitory concentration; *k*_a_, association rate constant; *k*_d_, dissociation rate constant; *K*_D_, equilibrium dissociation constant; mAb, monoclonal antibody; ND, not detected; RBD, receptor binding domain; SARS-CoV-2, severe acute respiratory syndrome coronavirus 2; SD, standard deviation; VOC, variant of concern.

### AZD7442 potently neutralizes SARS-CoV-2 variants of concern (VOCs) *in vitro*

The high binding affinities of the AZD7442 antibodies for the viral spike protein translated into potent SARS-CoV-2 neutralization activity. AZD8895, AZD1061 and AZD7442 potently neutralized the USA-WA1/2020 strain of SARS-CoV-2 (IC_50_ 9, 32, and 10 ng/mL, respectively) (**Fig. 1D**). Importantly, as a result of a combination of two mAbs with non-overlapping epitopes, AZD7442 retained potent neutralizing activity (fold-change IC_50_ <3.0) against SARS-CoV-2 Alpha, Beta, Gamma, and Delta VOCs compared with USA-WA/1/2020 or AUS/IC01/2020 reference strains in SARS-CoV-2 neutralization assays (**Fig. 1E**).

### AZD7442 exhibits extended half-life *in vivo* and reduced Fc effector functions *in vitro*

AZD7442 includes YTE and TM modifications in the antibody Fc regions. *In vitro*, AZD8895 and AZD1061 exhibited approximately 9-fold greater affinities for the neonatal Fc receptor (FcRn) than AZD8895-TM and AZD1061-TM (mAbs with TM but not YTE substitutions; *K*_D_ 272, 283, 2,400 and 2,360 nM, respectively) (**Fig. 2A**). This increased affinity for FcRn translated into extended half-lives (t_1/2_) for AZD8895 and AZD1061 in NHPs compared with mAbs without YTE modifications following intravenous (IV) administration of 600 mg/kg AZD7442. AZD1061 and AZD8895 had a median t_1/2_ of 19 days based on pharmacokinetic data collected over 8 weeks compared with t_1/2_ of 8–10 days for IgG in NHPs (*23*) (**Fig. 2B**). AZD8895 and AZD1061 at physiological serum concentrations demonstrated little or no binding to various FcγRs or C1q compared with AZD7442 antibodies with wild-type (WT) Fc (no YTE or TM substitutions; **Fig. 2C**). *In vitro* assays further confirmed that the AZD7442 antibodies displayed little or no Fc effector function, including antibody-dependent cellular phagocytosis (ADCP), antibody-dependent cellular cytotoxicity (ADCC), antibody-dependent complement deposition (ADCD), or antibody-dependent natural killer cell activation (ADNKA). AZD7442, AZD8895, and AZD1061 also did not mediate antibody-dependent enhancement of infection (**Fig. 2D** **and Fig. S2)**.

**Figure 2.**
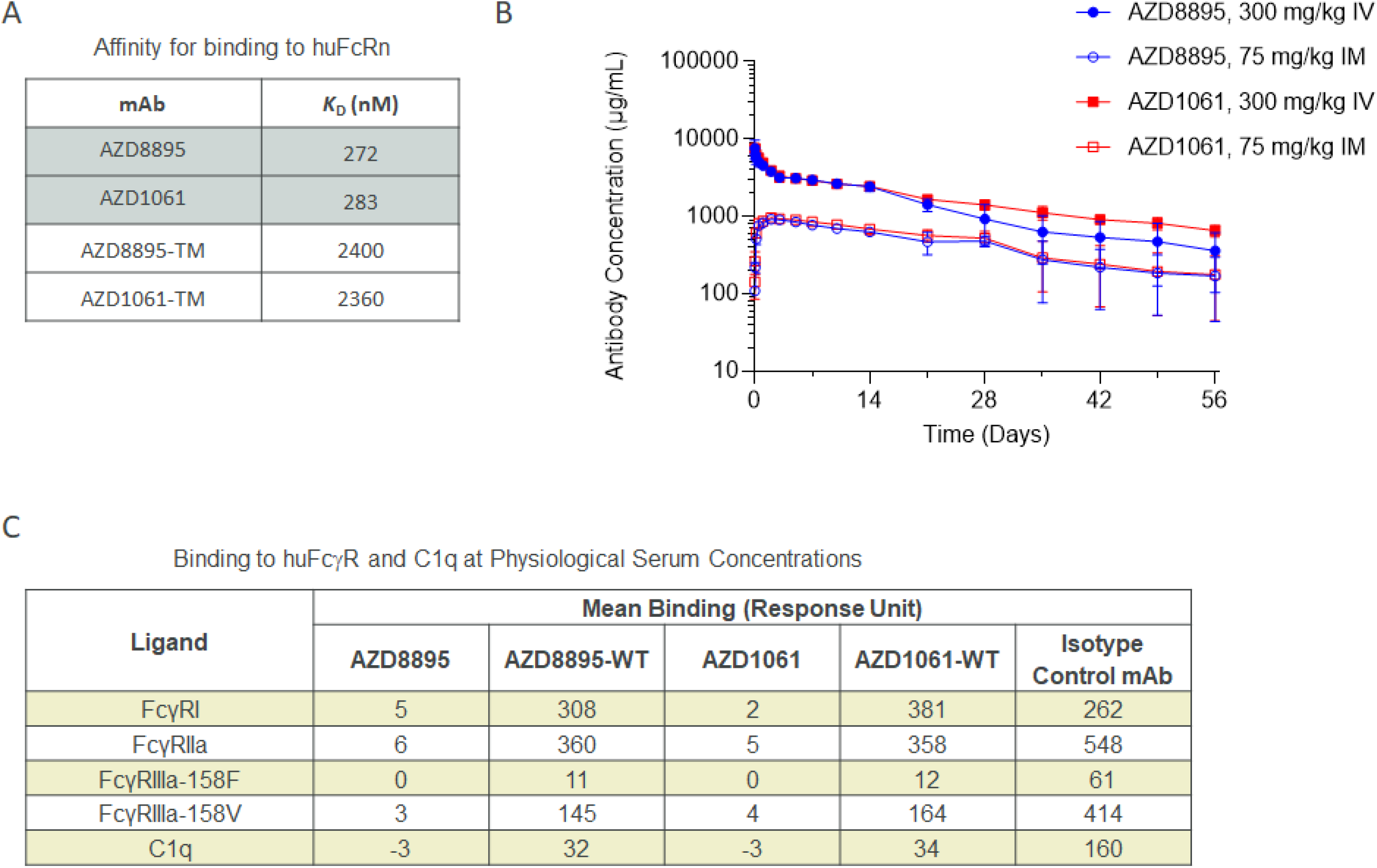
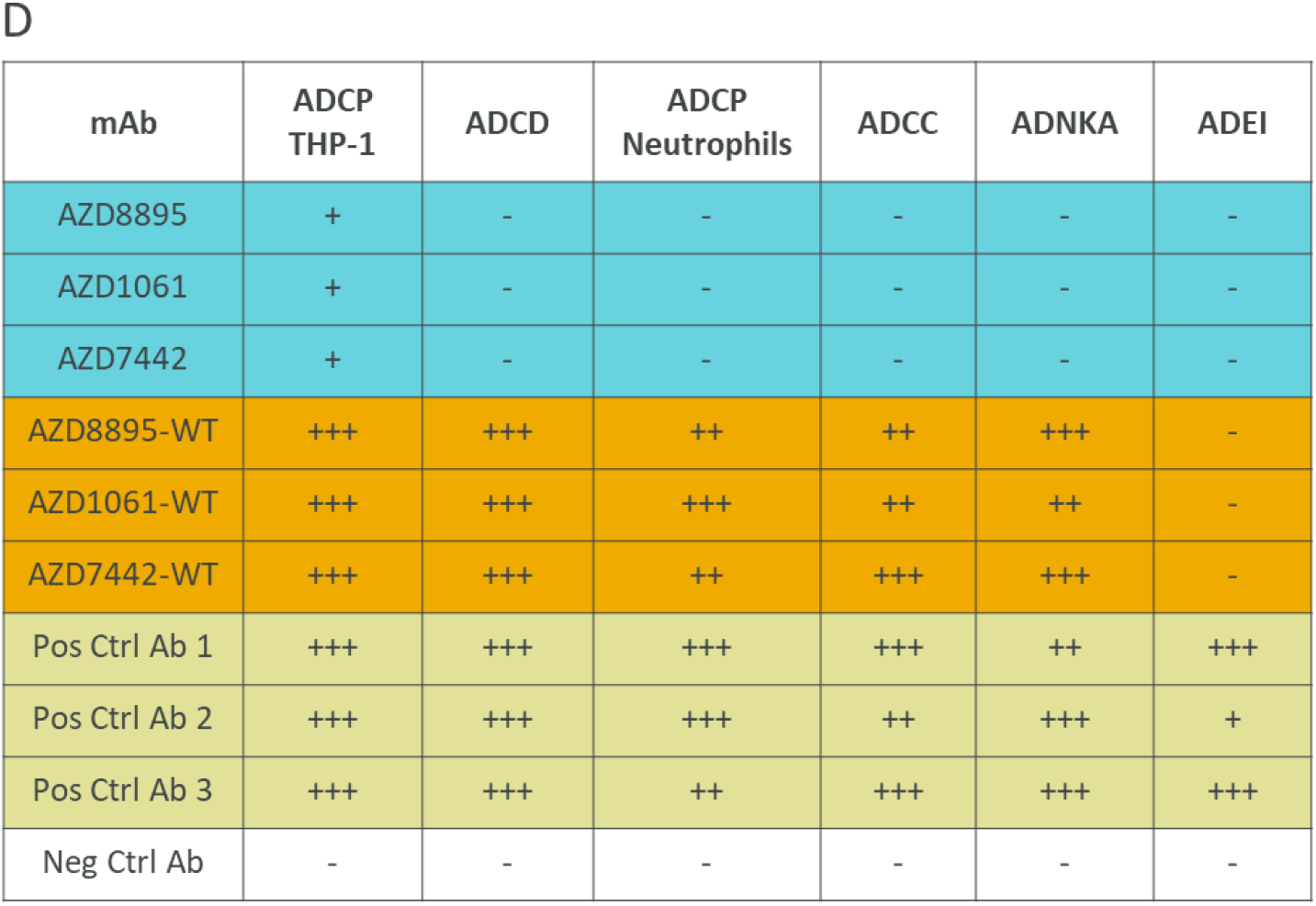
AZD7442 shows extended half-life and reduced Fc effector function *in vivo*. **A.** AZD8895 and AZD1061 binding affinities for human FcRn, measured by SPR with mAbs immobilized and titrated binding of huFcRn at pH 6.0. **B.** AZD8895 and AZD1061 exhibited extended *in vivo* serum half-lives in NHPs. Data shown are mean ± SD **C.** AZD8895 and AZD1061 exhibited reduced binding to FcγR and C1q at physiological serum mAb concentrations. Isotype control = R347-WT, an antibody to HIV glycoprotein gp120 with no TM or YTE modifications in the Fc. Binding response = mAb binding response / ligand binding to probe (background subtraction). **D.** AZD8895, AZD1061 and AZD7442 exhibited reduced Fc effector functions compared with WT mAbs. Ratings were based on normalized AUC values with –, +, ++ and +++ assigned to values <25%, ≥25-50%, ≥50-75%, and ≥75% AUC, respectively. Negative control is an antibody to Ebola virus glycoprotein with no TM or YTE modifications in the Fc; Positive controls are two antibodies to SARS-CoV-2 spike RBD with no TM or YTE modifications in the Fc (Pos. Ctrl 1 and 2) and their combination (Pos. Ctrl 3). Ab, antibody; ADCC, antibody-dependent cellular cytotoxicity; ADCD, antibody-dependent complement deposition; ADCP, antibody-dependent cellular phagocytosis; ADEI, antibody-dependent enhancement of infection; ADNKA, antibody-dependent natural killer cell activation; AUC, area under the concentration curve; Fc, fragment crystallizable; FcγR, Fc gamma receptor; FcRn, neonatal Fc receptor; IM, intramuscular; IV, intravenous; *K*_D_, equilibrium dissociation constant; mAb, monoclonal antibody; Neg Ctrl, negative control; Pos Ctrl, positive control; SD, standard deviation; SPR, surface plasmon resonance; TM, substitution L234F/L235E/P331S in the antibody Fc region; WT, wild-type antibody (no substitution in Fc region).

### AZD7442 prevents and treats SARS-CoV-2 infections in NHPs

AZD7442 was evaluated in NHP models of SARS-CoV-2 infection in prophylaxis or treatment settings in two separate studies; one in rhesus macaques and one in cynomolgus macaques. For each study, 3–4 NHPs were administered either a 40 mg/kg dose of isotype control antibody or AZD7442 at a dose ranging from 0.04 to 40 to mg/kg by IV infusion 3 days prior to viral challenge (prophylaxis) or a 40 mg/kg dose of AZD7442 24 hours after viral challenge (therapeutic analysis). To evaluate the contribution of Fc effector function to viral clearance, one group from each study received AZD7442-YTE (with YTE but not TM modification) as either prophylaxis (4 mg/kg, rhesus macaque study) or treatment (40 mg/kg, cynomolgus macaque study). In the prophylaxis studies, AZD7442 serum levels increased proportionally with dose across the 0.04 mg/kg to 40 mg/kg dose range; animals that received 40 mg/kg AZD7442 as treatment showed near maximal serum concentrations within 1 day of dosing **(Fig. S3)**. AZD7442 concentrations in the serum translated into high levels of serum neutralizing antibody titers in rhesus macaques and in cynomolgus macaques. Median pseudovirus neutralization (Neut_50_) titers of 4.5 and 6.4 log_10_ were measured for rhesus macaques given 4 and 40 mg/kg AZD7442, respectively **(Fig. S5A‒B)**; median SARS-CoV-2 plaque reduction neutralization test (PRNT_50_) titers of 3.4 and 4.6 log10 were measured for cynomolgus macaques administered 4 and 40 mg/kg AZD7442, respectively **(Fig. S5C‒D)**. Low neutralizing titers were detected in the isotype control-treated rhesus macaques beginning Day 10 post-infection consistent with the development of an adaptive humoral immune response (**Fig. S5A-B**).

Rhesus macaques were challenged on Day 0 with 10^5^ plaque forming units (PFU) of SARS-CoV-2 strain USA-WA1/2020 (**Fig. 3A**). SARS-CoV-2 viral subgenomic messenger RNA (sgmRNA) was measured by quantitative reverse-transcription polymerase chain reaction (qRT-PCR) in bronchoalveolar lavage (BAL) and nasal swab samples up to 14 days after virus challenge. In rhesus macaques treated prophylactically with isotype control, peak levels of viral sgmRNA were detected on Day 2 post-infection, with means of 4.67 log_10_ copies/mL (BAL) and 4.73 log_10_ copies/swab (nasal swab) of sgmRNA detected (**Fig. 3B‒C**). In contrast, sgmRNA was undetectable in the BAL samples of rhesus macaques receiving AZD7442 or AZD7442-YTE (**Figure 3B**); low levels of sgmRNA were detected transiently (Day 2 only) in nasal swab samples from two of four rhesus macaques receiving 4 mg/kg AZD7442 (3.32–3.60 log_10_ copies/swab) and from one of four that received 4 mg/kg AZD7442-YTE (4.95 log_10_ copies/swab) (**Fig. 3C**). The undetectable levels of sgmRNA in the BAL samples from rhesus macaques that received as little as 4 mg/kg AZD7442 (comparable to the human 300 mg dose) indicates that prophylactic administration of AZD7442 can protect against SARS-CoV-2 lower respiratory tract infection in rhesus macaques. The lack of Fc effector function and complement binding did not affect efficacy in this model as the non-TM (AZ7442-YTE) and the TM-modified AZD7442 showed similar *in vivo* efficacies (**Fig.3B** **and 3C**). Rhesus macaques treated with AZD7442 24 hours after inoculation showed a reduction in viral sgmRNA 24 hours later (day 2) in BAL (**Fig. 3D**) but not in nasal swabs as compared to isotype control-treated NHPs (**Fig. 3E**). AZD7442 administration resulted in accelerated viral clearance as detected in both BAL and nasal swabs within 4 and 7 days post-infection, respectively. In comparison, NHPs that received the isotype mAb did not fully clear the virus until Day 10 post-infection (Fig. 3D‒E).

**Figure 3.**
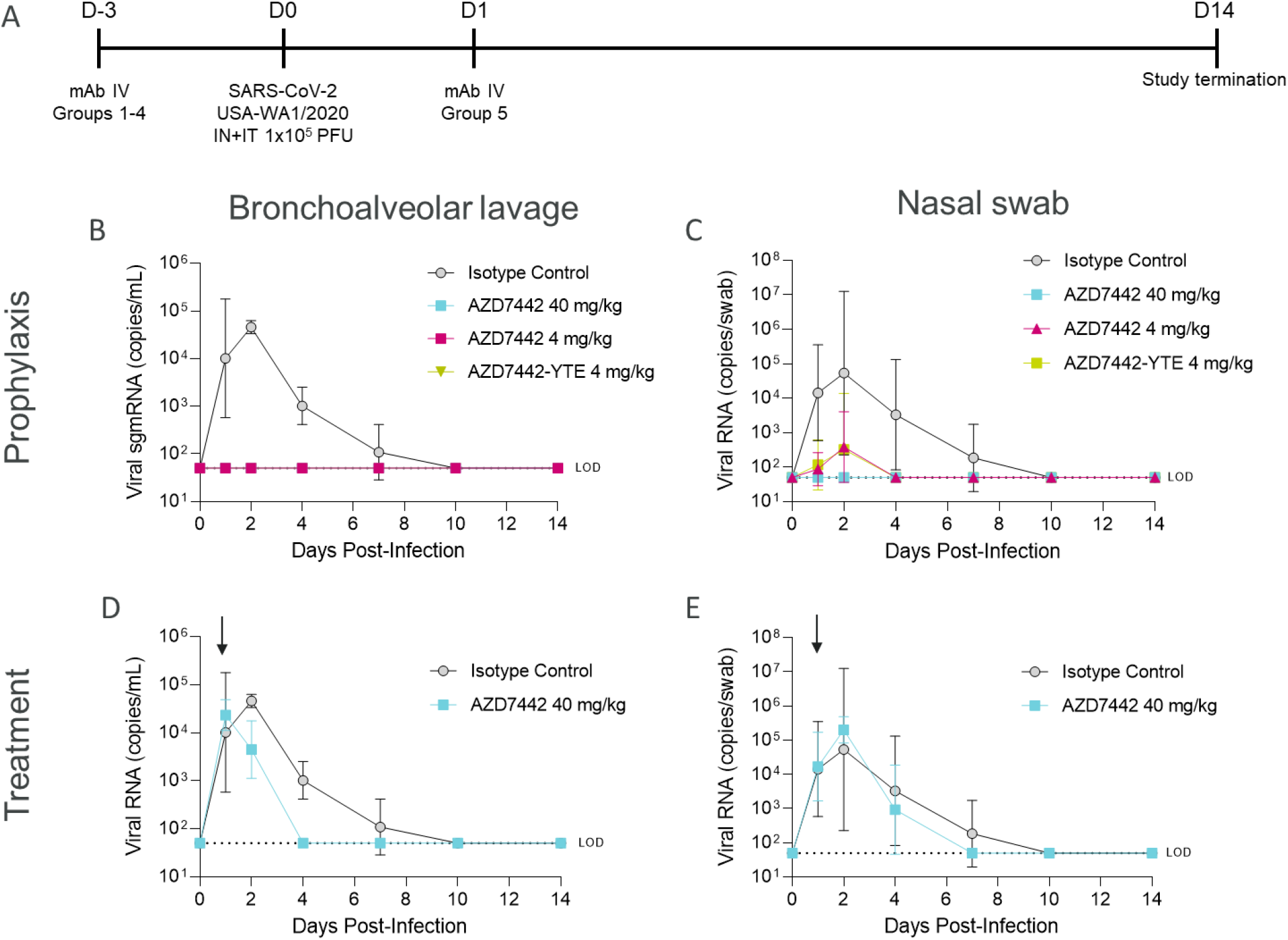
AZD7442 administration protects rhesus macaques against SARS-CoV-2 infection in prophylaxis or treatment settings. **A.** Timeline of the in vivo SARS-CoV-2 challenge study. Six-year-old rhesus macaques weighing ∼3.6–7.3 kg in prophylaxis groups 1, 2, 3, and 4 (n=3 for Groups 1 and 2; n=4 for Groups 3 and 4) received IV infusions of 40 mg/kg isotype control mAb R347-TM-YTE, 40 mg/kg AZD7442, 4 mg/kg AZD7442, or 4 mg/kg AZD7442-YTE, respectively, 3 days prior to challenge. Rhesus macaques in treatment group 5 (n=4) received an IV infusion of 40 mg/kg AZD7442 1 day after challenge. Rhesus macaques were challenged with 10^5^ PFU of SARS-CoV-2, split between IT and IN delivery on Day 0. BAL and nasal swab samples were collected at days 0, 1, 2, 4, 7, 10, and 14. One rhesus macaque from each of Groups 3, 4, and 5 was euthanized on Day 2 for histopathology analyses (data not shown as minimal inflammation was observed on Day 2 rendering results inconclusive). **B.** Geometric mean ± SD viral burden in BAL samples from rhesus macaques receiving isotype control mAb, AZD7442 or AZD7442-YTE as prophylaxis 3 days prior to SARS CoV-2 challenge **C.** Geometric mean ± SD viral burden in nasal swab samples from rhesus macaques receiving isotype control mAb, AZD7442 or AZD7442-YTE as prophylaxis 3 days prior to SARS CoV-2 challenge **D.** Geometric mean ± SD viral burden in BAL samples from rhesus macaques receiving isotype control mAb, AZD7442 or AZD7442-YTE as treatment 1 day after SARS CoV-2 infection **E.** Geometric mean ± SD viral burden in nasal swab samples from rhesus macaques receiving isotype control mAb, AZD7442 or AZD7442-YTE as treatment 1 day after SARS CoV-2 infection BAL, bronchoalveolar lavage; D, day; IN, intranasal; IT, intratracheal; IV, intravenous; LOD, limit of detection; mAb, monoclonal antibody; PFU, plaque forming unit; RNA, ribonucleic acid; sgmRNA, subgenomic messenger RNA; SARS-CoV-2, severe acute respiratory syndrome coronavirus 2; SD, standard deviation.

In the second study, cynomolgus macaques were challenged on Day 0 with 10^5^ tissue culture infection dose (TCID_50_) of SARS-CoV-2 strain USA-WA1/2020 (**Fig. 4A**). SARS-CoV-2 burden was measured in BAL and nasal swab samples up to 5 days after virus challenge. Prophylactic AZD7442 administration demonstrated dose-dependent reduction of infectious virus titers in BAL samples (**Fig. 4B**), and reduction in viral sgmRNA concentrations in BAL and nasal swab samples **(Fig. S4A and S4B)** compared with isotype control antibody. Importantly, the 4 mg/kg dose (comparable to the human 300 mg dose) was fully protective in both NHP studies. Also consistent with observations from the rhesus macaque study, faster virus clearance was observed in cynomolgus macaques administered 40 mg/kg AZD7442 therapeutically compared with isotype control (**Figure 4C**). AZD7442 treatment reduced viral titer similarly to AZD7442-YTE (no TM modification) at an equivalent dose, suggesting that Fc effector function and/or complement binding are not required for treating SARS-CoV-2 infection.

**Figure 4.**
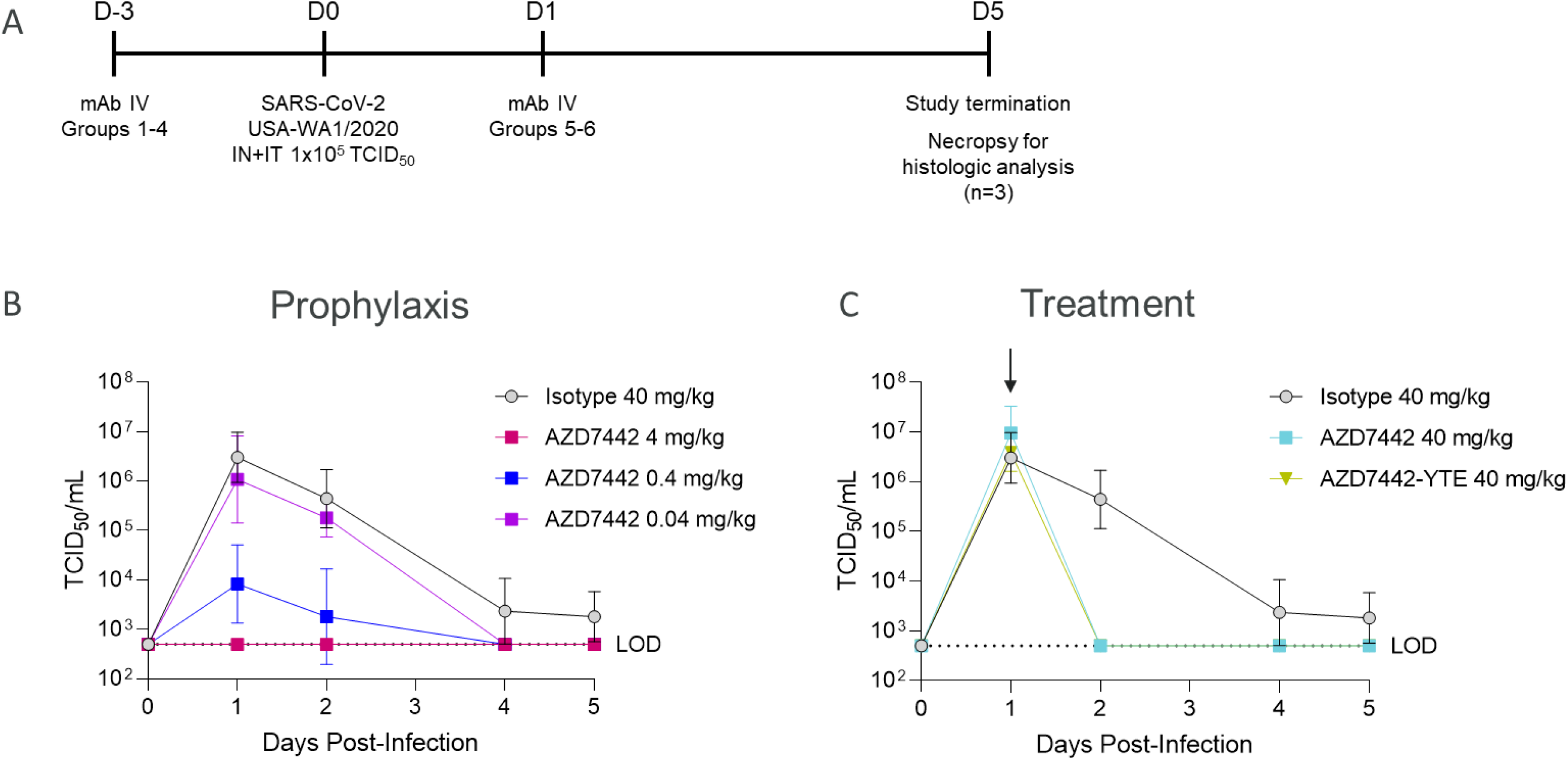
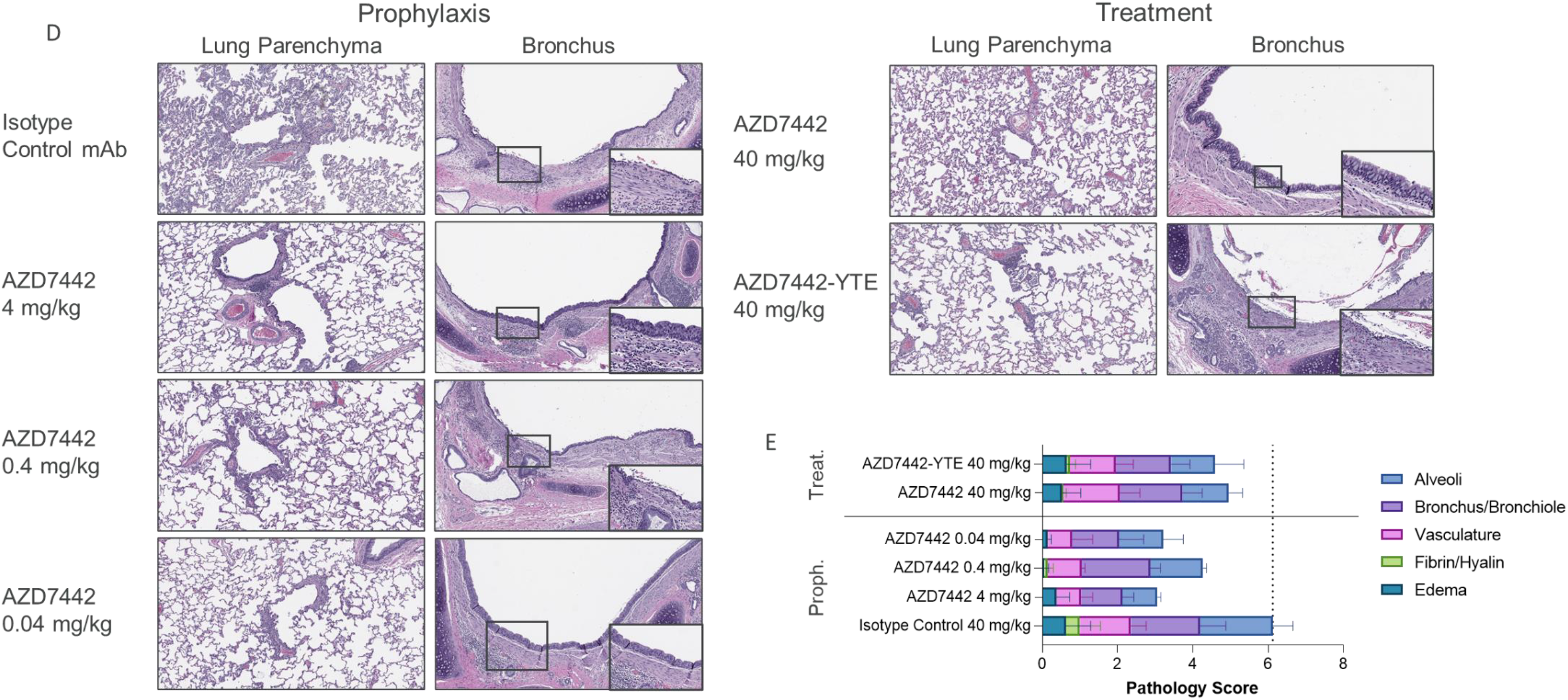
AZD7442 administration protects cynomolgus macaques against SARS-CoV-2 infection and associated lung immune pathology in prophylaxis and treatment. **A.** Timeline of the *in vivo* SARS-CoV-2 challenge study. 6-years-old cynomolgus macaques weighing ∼3.2–6.3 kg in prophylaxis Groups 1, 2, 3, and 4 (all groups n=3), received IV infusions of 40 mg/kg isotope control mAb R347-TM-YTE, 40 mg/kg AZD7442, 4 mg/kg AZD7442, or 4 mg/kg AZD7442-YTE, respectively, 3 days prior to challenge. Cynomolgus macaques in treatment Group 5 (n=3) received an IV infusion of 40 mg/kg AZD7442 1 day after challenge. Cynomolgus macaques were challenged with 10^5^ TCID_50_ of SARS-CoV-2, split between intratracheal and intranasal delivery on Day 0. BAL and nasal swab samples were collected at days 0, 1, 2, 4, 7, 10, and 14. Serum was collected on days -3, 0, 1, 2, 4, 7, 10 and 14. One cynomolgus macaque from each of Groups 3, 4, and 5 was euthanized on Day 2 for histopathology analyses. **B.** Mean ± SD viral burden in BAL samples from cynomolgus macaques receiving isotype control mAb or AZD7442 as prophylaxis 3 days prior to SARS CoV-2 challenge. **C.** Mean ± SD viral burden in BAL samples from control cynomolgus macaques receiving isotype control mAb, or cynomolgus macaques receiving AZD7442 or AZD7442-YTE as treatment 1 day after SARS CoV-2 infection. **D.** Lung histology following AZD7442 administration in cynomolgus macaques. Hematoxylin/eosin-stained lung parenchyma and bronchus from a representative animal (10x magnification). Inset shows 20x magnified image of bronchus. **E.** Mean + SD pathology score assigned by board-certified veterinary pathologist based on histologic findings on 8 lung sections per animal (n=3). The scores for one animal from the AZD7442 4 mg/kg prophylaxis dose group was excluded from analysis due to evidence of foreign material (plant matter) in multiple sections and observed inflammation inconsistent with SARS-CoV-2 infection BAL, bronchoalveolar lavage; D, day; IN, intranasal; IT, intratracheal; IV, intravenous; LOD, limit of detection; mAb, monoclonal antibody; proph, prophylaxis; SARS-CoV-2, severe acute respiratory syndrome coronavirus 2; SD, standard deviation; TCID_50_, tissue culture infection dose; treat, treatment.

Lung sections from control cynomolgus macaques showed histologic changes consistent with SARS-CoV-2 infection (**Fig. 4D**). In sections of lung parenchyma, there was mild to marked perivascular cuffing of medium-caliber blood vessels by lymphocytes, alveolar wall thickening, and mixed inflammatory cell infiltrates comprising lymphocytes, neutrophils, and macrophages within alveolar spaces. There was mild to marked bronchial/bronchiolar inflammation with loss and/or blunting of bronchial/bronchiolar epithelium and infiltrates of lymphocytes and neutrophils within the lamina propria and submucosa. Although there were some variability between sections consistent with focal SARS-CoV-2 infection, lung sections from animals administered AZD7442 either prophylactically or therapeutically showed an overall reduction in inflammation compared with those treated with isotype control. The 4 mg/kg prophylaxis group demonstrated minimal to mild histological changes in the lung parenchyma and bronchial epithelium compared with all other AZD7442 doses tested in both prophylaxis and treatment settings in NHPs. Pathology scores were similarly reduced for animals that received 40 mg/kg AZD7442 or AZD7442-YTE as treatment (**Fig. 4E**), suggesting that AZD7442 with TM modification provides equivalent protection against SARS-CoV-2-induced lung injury to AZD7442 without TM modification.

Taken together, these two NHP studies indicate that AZD7442 administration prophylactically protects NHPs against SARS-CoV-2 infection, and therapeutically accelerates viral clearance. Importantly, AZD7442 reduced pulmonary inflammation and protected animals against alveolar damage associated with SARS-CoV-2 infection, suggesting AZD7442 can provide clinical benefit in both prevention and treatment settings.

### AZD7442 exhibits extended half-life and provides high anti-SARS-CoV-2 neutralizing antibody levels in healthy adult participants

The pharmacokinetic characteristics and transudation of AZD7442 to the nasal mucosae were evaluated in a randomized, double-blind, placebo-controlled, Phase I study of 60 healthy adults (NCT04507256). This study is ongoing and the data presented herein are preliminary. Full results will be published in due course. Participants in the active arm received 300 mg of IM AZD7442 (150 mg of each mAb administered sequentially), IV AZD7442 at doses of 300 mg, 1,000 mg or 3,000 mg (150 mg, 500 mg or 1,500 mg, respectively of each mAb administered sequentially), or 3,000 mg AZD7442 co-administered by IV (1,500 mg AZD8895 plus 1,500 mg AZD1061); all groups n=10.

Serum concentrations of AZD8895 and AZD1061 were measured up to 9 months post-dose and confirmed the extended t_1/2_ of approximately 90 days for both antibodies in each dose cohort following either IV or IM administration (**Fig. 5A**). After a single 300 mg IM dose, the geometric mean C_max_ of AZD8895 (16.5 μg/mL) and AZD1061 (15.3 μg/mL) were similar and reached at a median T_max_ of 14 days.

**Figure 5.**
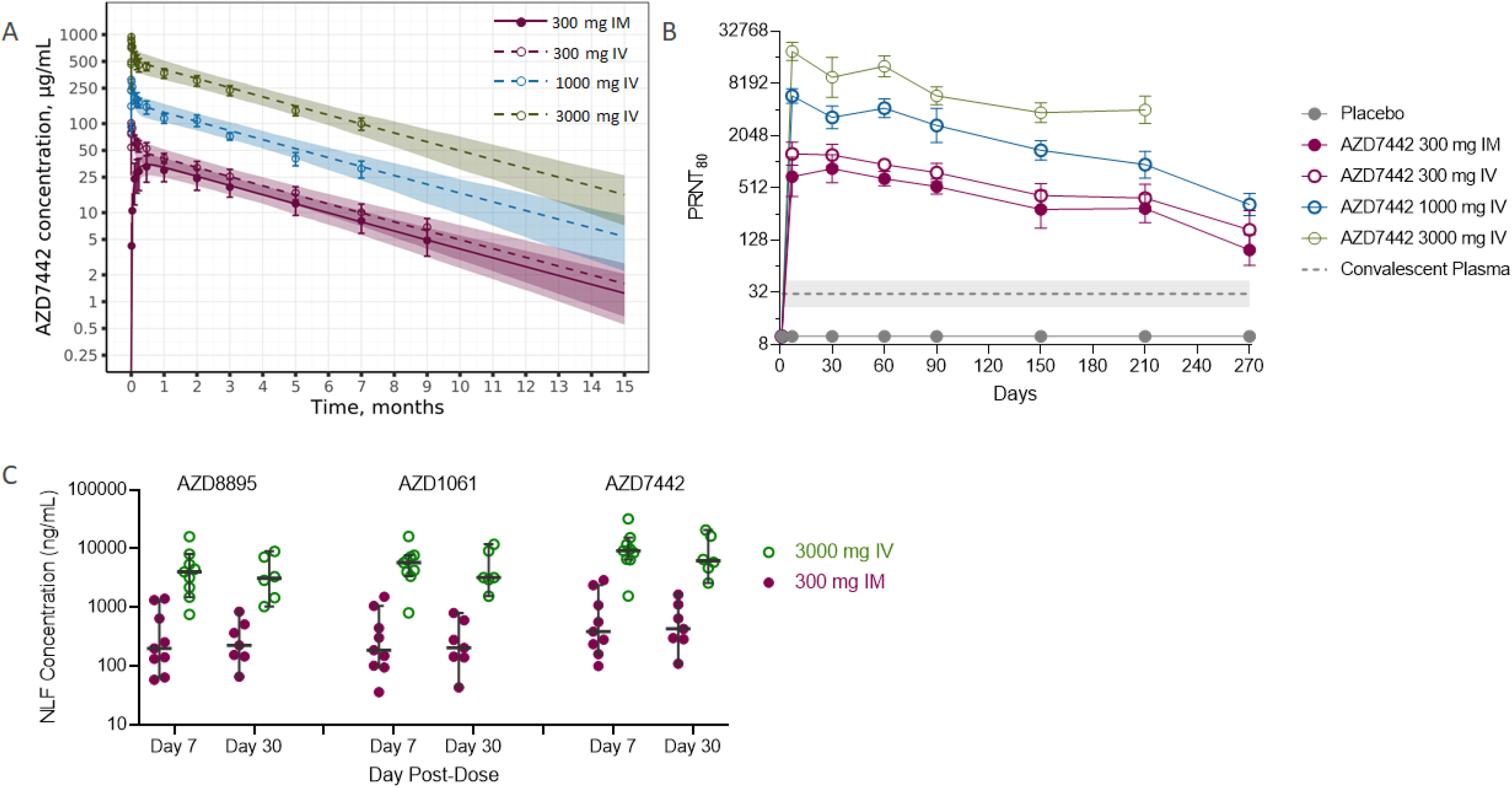
AZD7442 exhibits extended half-life, confers high anti-SARS-CoV-2 neutralizing antibody levels and transudates to the mucosal epithelium in healthy adult participants. **A.** Symbols are observed mean ± SD serum concentrations of AZD7442 over 9 months following single IM or IV administration of AZD7442 in healthy participants, and lines represent the predicted mean with shaded area representing the 90% prediction interval up to 15 months. **B.** Geometric mean neutralizing antibody titers against SARS-CoV-2 over 9 months following single IM or IV doses in healthy adult participants; data represent geometric mean PRNT_80_ titer ± SD for placebo or AZD7442 and GMT ± 95% CI for convalescent plasma samples. **C.** Concentrations of AZD8895, AZD1061 and AZD7442 (AZD8895 + AZD1061) in the NLF following 300 mg IM or 3000 mg IV doses of AZD7442. Graph shows individual and median concentration ± 95% confidence interval. IM, intramuscular; IV, intravenous; NLF, nasal lining fluid; PRNT_50_, median plaque reduction neutralization titer; GMT, geometric mean neutralizing antibody titer; SARS-CoV-2, severe acute respiratory syndrome coronavirus 2; SD, standard deviation.

SARS-CoV-2 neutralizing antibody titers in sera conferred by AZD7442 were considerably higher than titers associated with convalescent plasma (**Fig. 5B**). Geometric mean neutralization titers (GMT) afforded by AZD7442 300 mg IM and IV were ∼22 and 41-fold higher, respectively, 7 days after dosing than those of convalescent sera. At Day 270 post dose, GMT following AZD7442 administration remained 3-fold higher than the GMT in convalescent plasma samples (n=28). For the 300 mg IM dose, median AZD8895 and AZD10161 nasal lining fluid (NLF) concentrations were ∼200 ng/mL through 30 days post-dose (**Figure 5C**). This correlates to 1–2% of AZD8895 and AZD1061 levels in the sera, and importantly, suggest high levels of neutralizing antibodies at a key site of SARS-CoV-2 infection.

Extrapolating serum and nasal AZD7442 concentrations and anti-SARS-CoV-2 neutralizing antibody titers to 15 months post dose using an estimated t_1/2_ of ∼90 days suggest that a 300 mg intramuscular dose of AZD7442 could provide protection against COVID-19 for up to 12 months (**Figure 5A**).

## Discussion

Antibody combinations comprising mAbs targeting distinct epitopes on the SARS-CoV-2 spike protein provide a higher threshold against virus escape from neutralization than single antibodies (*24*), as the virus would have to acquire mutations at both epitopes to escape neutralization. Structural analysis of the mAbs comprising AZD7442 (AZD8895 and AZD1061) showed that they simultaneously bind the RBD at distinct, non-overlapping epitopes at the RBD-ACE2 interface (*24*). These data reinforce prior analyses of the AZD8895 and AZD1061 progenitor antibodies, COV-2196 and COV-2130, which demonstrated that they neutralize SARS-CoV-2 by blocking binding to ACE2 (*19, 25*). Furthermore, we show that a different Fc domain containing TM and YTE modifications did not affect binding to the RBD.

Mutations in the spike protein have been shown to increase viral infectivity and transmissibility (*26, 27*) and/or threaten the effectiveness of vaccines and therapeutics by providing escape from neutralizing antibodies (*28–31*). Monoclonal antibody combinations that target distinct SARS-CoV-2 spike protein epitopes have demonstrated a higher threshold against virus escape than the individual mAbs (*28, 32*). AZD8895 and AZD1061 individually or in combination potently neutralized the USA-WA1/2020 strain of SARS-CoV-2. Importantly, we demonstrated in this study that AZD7442 retains potency to neutralize all SARS-CoV-2 VOC, including those with spike mutations that confer reduced susceptibility to AZD8895 or AZD1061 individually. Taken together, the data presented here and in other studies confirm that AZD7442 retains activity against all known VOCs, including the Alpha (*24, 28-30, 33*), Beta (*24, 29-31, 33*), Gamma (*28, 30, 34*) and importantly, Delta (*24, 35*).

In some virus infections, antibody-enhanced disease mediated by Fc engagement of immune cells and immune complex formation leading to enhanced immune activation has been proposed as a mechanism of exacerbation of immunopathology and inflammation (*36*). The TM modification in the Fc region was therefore introduced to reduce this potential risk by preventing FcγR and complement binding. This study confirmed that the introduction of the TM modification to AZD7442 abrogated Fc effector function without affecting neutralization activity. While Fc effector function has been shown to enhance viral clearance by some mAbs in mouse models of SARS-CoV-2 infection (*37, 38*), SARS-CoV-2 clearance was not affected by the TM substitution in the NHP studies of AZD7442 reported here. These findings support the continued clinical evaluation of this TM-modified mAb combination for the prevention and treatment of COVID-19.

AZD7442 was also modified to include the half-life extending YTE substitutions (*20, 21*). YTE-modified mAbs are under clinical evaluation in other infectious diseases, including for immunoprophylaxis against respiratory syncytial virus in infants (*39*). AZD8895 and AZD1061 have higher affinities for human FcRn at low endosomal pH than antibodies without the YTE modification, which translated into extended half-lives in NHPs and humans, consistent with other studies of YTE-enhanced mAbs (*20, 40, 41*).

At the 4 mg/kg dose, prophylactic AZD7442 administration prevented lower respiratory tract infection in NHPs as measured by viral sgmRNA or infectious virus titer in BAL samples. This is comparable with the prophylactic effect observed with other mAbs evaluated in similar NHP models (*42, 43*). Viral sgmRNA was also largely undetectable in nasal swabs at this dose, suggesting that AZD7442 administration can prevent SARS-CoV-2 infection and/or virus shedding from the upper respiratory tract. The therapeutic administration of 40 mg/kg AZD7442 accelerated viral clearance as measured by viral sgmRNA levels or infectious virus titers in BAL and nasal swab samples. This finding contrasts with studies of remdesivir in rhesus macaques, where remdesivir reduced viral titers in BAL samples but not in the upper respiratory tract (*44*). Histologically, alveolar and bronchial sections of lungs from NHPs administered AZD7442 prophylactically or therapeutically all displayed reduced inflammation and pulmonary tissue damage associated with SARS-CoV-2 infection. Importantly, NHPs treated with the 4 mg/kg dose of AZD7442 exhibited serum antibody concentrations similar to those observed in indivduals in the Phase 1 study after they received a 300 mg IM dose of AZD7442, demonstrating the clinical relevance and translatability of the doses evaluated in these NHP models of SARS-CoV-2 infection.

While NHP models do not completely mimic severe COVID-19 disease in humans, these findings support the further clinical evaluation of AZD7442 in COVID-19 prevention and treatment settings. The 4 mg/kg AZD7442 IV dose that demonstrated prevention of SARS-CoV-2 infection in NHPs resulted in serum concentrations comparable to the levels achieved with a 300 mg AZD7442 IM dose delivered to healthy adult participants. The 300 mg AZD7442 IM dose conferred an anti-SARS-CoV-2 neutralizing antibody GMT at Day 270 post-dose that was ∼3-fold greater than those detected in convalescent plasma. In addition, pharmacologically active concentrations of AZD7442 in the nasal mucosa at 7 and 30 days post-dose were on average 40-fold above the IC_50_ of AZD7442. These data support the potential of a 300 mg IM dose of AZD7442 to provide protection against COVID-19 for up to 12 months.

In conclusion, AZD7442 demonstrated potent *in vitro* neutralization against SARS-CoV-2 VOCs, *in vivo* efficacy in both prevention and treatment settings of SARS-CoV-2 infection in NHPs, and extended half-life in NHPs and healthy adult participants. Pharmacokinetic predictions support that AZD7442 could provide up to 12 months of protection from COVID-19. AZD7442 therefore has the potential to provide almost immediate protection in unvaccinated individuals or bolster the immune response of individuals who respond sub-optimally to vaccination, such as those with hematologic malignancies (*7*) or solid organ transplant recipients (*6*). Accelerated viral clearance and reduced immunopathology in NHPs support potential use of AZD7442 as treatment for COVID-19 to prevent disease progression. The results of the Phase 2 and 3 clinical trials evaluating AZD7442 in key settings of pre-exposure prophylaxis (NCT04625725), out-patient treatment (NCT04723394 and NCT04518410), and in-patient treatment (NCT04501978 and NCT04315948) will determine if AZD7442 is efficacious in preventing and treating COVID-19, the results of which are eagerly anticipated.

## Materials and Methods

Additional materials and methods are provided in the Supplementary Materials describing: hydrogen-deuterium exchange mass spectrometry; protein expression and purification; binding assays; virus neutralization assays; and Fc effector function analyses.

### AZD7442 Pharmacokinetic Study in NHPs

#### Group designation and study design

The pharmacokinetic analyses were part of an 8-week GLP toxicity study conducted at Charles River Laboratories. Cynomolgus monkeys were received from Orient BioResource (Alice, Tx, USA) and were between 2.2 and 5.1 years-old; males weighing between 1.9 and 2.6 kg and females weighing between 2.2 and 2.5 kg at the time of dosing. Animals were randomized and assigned to groups using a computer-based procedure prior to transfer to study; males and females were randomized separately. AZD7442 was administered to cynomolgus monkeys by a IV infusion at 600 mg/kg (n = 10) or by a IM injection at 300 mg/kg (n = 4); control animals were administered vehicle alone at volumes equivalent to AZD7442 by a single IV infusion (n = 12) or a single IM injection (n = 4). For the IV administered animals, all toxicology/safety assessment endpoints were evaluated at Day 15 (n = 9 control animals and n = 6 AZD7442 animals) and Day 57 (n = 4 for control and AZD7442 animals) necropsy timepoints. The IM animals had only general in-life procedures, observations and measurements collected.

IV infusion was via a suitable peripheral vein using an infusion pump connected to a temporary indwelling catheter once for 15 minutes. AZD8895 and AZD1061 were administered via separate 15 minute infusions, for a total infusion time of 30 minutes. A 1 mL saline flush was administered following each 15 minute infusion. IM injections to the anterior thigh was performed on temporarily restrained but not sedated animals. AZD8895 and AZD1061 were administered separately via 2 injections, 1 injection per thigh. In-life measurements included post-dose and detailed clinical observations, injection site dermal scoring, body weights, food consumption, veterinary physical examinations, ophthalmic examinations, electrocardiography exams, neurologic examination, blood pressure and heart rate, respiratory rate and body temperature. Blood was collected by venipuncture at various times before AZD7442 administration and up to 8 weeks post dose to measure AZD8895 and AZD1061 serum concentations and *ex vivo* pharmacodynamic evaluations. At study termination, animals were euthanized by IV injection of a commercially available veterinary euthanasia solution, followed by exsanguination.

### SARS-CoV-2 Challenge Studies in NHPs

#### Group designation and study design

Animal studies (IACUC protocol no. 20-035 and 21-018P) were approved by the Institutional Animal Care and Use Committee and conduced at BIOQUAL, Inc. (Rockville, MD, USA) in adherence of the following standards of the Association for Assessment and Accreditation of Laboratory Animal Care: the 8^th^ edition of the *Guide for the Care and Use of Laboratory Animals*; the *Animal Welfare Act*; and the *2015 reprint of the Public Health Service Policy on Human Care and Use of Laboratory Animals*. The total number of animals used, group sizes and number of groups were considered the minimum required to properly characterize the effects of AZD7442, AZD7442-YTE and the isotope control mAb R347-TM-YTE, and the study was designed so that it did not need an unnecessary number of animals to accomplish its objectives.

NHPs were quarantined and allowed to acclimatize for at least 7 days at BIOQUAL, Inc. Upon initiation of SARS-CoV-2 challenge procedures, the NHPs were housed in HEPA-filtered microisolator caging units. Cage-side observations were performed at least twice daily. Physical assessments of anesthetized NHPs included heart rate, body weight and rectal temperature at the time of sedation. No NHPs were excluded after none showed signs of illness or poor health. Veterinary care was provided by full-time veterinarians. NHPs received a primate diet (Purina, Monkey Diet Jumbo) *ad libitum* and were provided environmental enrichment by BIOQUAL, Inc, in accordance with the requirements of the Office of Laboratory Animal Welfare, the USA Animal Welfare Act, and the Guide for the Care and Use of Laboratory Animals.

For the first NHP study, 14 rhesus macaques (*Macaca mulatta*) of Indian origin, aged 5– 6 years and weighing between 3.6 and 7.3 kg, received prophylactic 10 mL IV infusions of the following mAbs 3 days prior to SARS-CoV-2 challenge: 40 mg/kg isotype control mAb R347-TM-YTE (Group 1; 2 male, 1 female); 40 mg/kg AZD7442 (Group 2; 2 male, 1 female); 4 mg/kg AZD7442 (Group 3; 2 male, 2 female); or 4 mg/kg AZD7442-YTE (Group 4; 3 male, 1 female). Four additional rhesus macaques weighing between 4.6 and 6.6 kg received a therapeutic 10 mL IV infusion of 40 mg/kg AZD7442 one day after SARS-CoV-2 challenge (Group 5; 2 male, 2 female).

In the second NHP study, 18 cynomolgus macaques (*Macaca fascicularis*) of Cambodian origin, aged 4–5 years old and weighing between 3.2 and 5.6 kg, received prophylactic 10 mL IV infusions of the following mAbs three days prior to SARS-CoV-2 challenge: 40 mg/kg isotype control mAb R347-TM-YTE (Group 1; 1 male, 2 female); 4 mg/kg AZD7442 (Group 2; 1 male, 2 female); 0.4 mg/kg AZD7442 (Group 3; 1 male, 2 female); or 0.04 mg/kg AZD7442 (Group 4; 2 male, 1 female). Six additional cynomolgus macaques weighing between 3.4 and 6.3 kg received therapeutic 10 mL IV infusions of 40 mg/kg AZD7442 (Group 5; 2 male, 1 female) or 40 mg/kg AZD7442-YTE (Group 6; 2 male, 1 female) 1 day after SARS-CoV-2 challenge.

All NHPs were infected with SARS-CoV-2 strain USA-WA1/2020 on Day 0. Serum samples were collected for human IgG concentration and neutralization titer measurements. BAL samples and nasal swabs were collected for virologic analyses. One rhesus macaque from each of Groups 3–5 was euthanized on Day 2 post-infection for histopathology analyses. Cynomolgus macaques from all groups were euthanized on Day 5 post-infection for histopathology analyses. The design for each study is summarized in **Fig. 3A** and **Fig. 4A**.

#### SARS-CoV-2 challenge procedures

All NHPs were challenged on Day 0 with 10^5^ PFU (rhesus macaques) or 10^5^ TCID_50_ (cynomolgus macaques) SARS-CoV-2 variant USA-WA1/2020. Rhesus macaques were challenged with an in-house generated stock, lot no. 022320-1100, expanded in Vero E6 cells from BEI Resources (Manassas, VA, USA), cat. no. NR-52281, lot no. 70033175 (*45*). Cynomolgus macaques were challenged with a stock, obtained from BEI Resources, cat. no. NR-53872, lot 70040665, 6.9 × 10^4^ TCID_50_ per mL in Vero E6 cells. Virus inoculum was prepared on the day of administration at BIOQUAL, Inc. by research associates. SARS-CoV-2 was administered to NHPs under ketamine sedation, with the virus inoculum split between intratracheal and intranasal routes.

#### Sample collection

Blood was collected under anesthesia by femoral venipuncture into BD Vacutainer SST gel tubes. BAL samples were collected under anesthesia via rubber feeding tubes inserted into the trachea. Nasal swab samples were collected under anesthesia using flocked swabs (COPAN Diagnostics Inc., Carlsbad, CA, USA).

#### Quantitation of SARS-CoV-2 genomic RNA and sub-genomic mRNA by qRT-PCR

RNA was extracted from BAL and nasal swab samples using a QIAcube HT (Qiagen, Germantown, MD, USA) and the Cador pathogen HT kit and eluted in nuclease-free water. RNA was reverse transcribed using superscript VILO (Thermo Fisher Scientific, Waltham, MA, USA) and tested in duplicate with QuantStudio 6 and 7 Flex RTPCR System (Thermo Fisher Scientific). Viral titers were calculated to give copies/mL using a viral RNA standard curve, using the primers and probes that target the SARS-CoV-2 E gene sub-genomic mRNA (sgmRNA). To generate a standard curve for the SARS-CoV-2 E gene sgmRNA assay, the SARS-CoV-2 E gene sgmRNA was cloned into a pcDNA3.1 expression plasmid. This insert was transcribed using the AmpliCap-Max-T7 High Yield Message Marker Kit (Cellscript, Madison, WI, USA) to obtain RNA (LOD 50 copies/mL).

#### Quantitation of human IgG in NHP serum samples collected in the SARS-CoV-2 challenge studies

The concentrations of AZ7442, AZD7442-YTE, and control R347-TM-YTE in NHP serum samples were determined by ELISA for human IgG against a known standard curve. The absorbance at 450 nm was recorded using a VersaMax or Omega microplate reader. Background-subtracted absorbance values for each standard was fit to a non-linear, sigmoidal (4PL) curve using GraphPad Prism, and the ensuing curve used to calculate concentrations of human IgG in each serum sample.

#### SARS-CoV-2 Pseudovirus neutralization assay

A pseudovirus neutralization assay (*46*) was used to quantify neutralizing antibody titers in rhesus macaque serum samples.

#### Histology

Lungs were removed immediately after euthanasia. NHP lung tissues were fixed in 10% neutral-buffered formalin and submitted in 70% ethanol to the histology laboratory at AstraZeneca (Gaithersburg, MD, USA). Paraffin-embedded lung samples were sectioned at 4 μM and stained with hematoxylin and eosin for evaluation by a board-certified veterinary pathologist.

### Phase I Clinical Study

#### Clinical study conduct

This is an ongoing Phase I, randomized, double-blind placebo-controlled, dose-escalation study of the safety, tolerability, and pharmacokinetics of AZD7442 in healthy adult participants with no prior history of COVID-19 and no prior receipt of a COVID-19 vaccine (NCT04507256). Participants, investigators, clinical staff, and the study monitor were all blinded from the assigned intervention. The study was conducted in compliance with the ethical principles originating in or derived from the Declaration of Helsinki and in compliance with the International Confederation on Harmonization Good Clinical Practice Guidelines. All participants provided written informed consent before entering the study. The study protocol and informed consent documentation were reviewed and approved by the study site institutional review board.

#### Clinical sampling

Sixty healthy participants aged 18–55 years at a single site in the United Kingdom were randomized 5:1 to one of five dose cohorts to receive one of four AZD7442 doses (300 mg IM, 300 mg IV, 1,000 mg IV or 3,000 mg IV, each mAb administered sequentially at 50% of the total AZD7442 dose; 3,000 mg IV co-administration of AZD8895 and AZD1061 as a single mixed infusion) or placebo. To determine the pharmacokinetics of AZD7442, serum samples were collected and analyzed pre-dose (all cohorts), during infusion and after infusion (IV cohorts only), at 8 hours post-dose and 1, 3, 5, 7, 14, 30, 60, 90, 150, and 210 days post-dose (all cohorts), and 270 days post-dose (300 mg dosing cohorts only). To determine the SARS-CoV-2 neutralizing activity of AZD7442, serum samples were collected and tested pre-dose and 7, 30, 60, 90, 150, and 210 days post-dose (all cohorts) and 270 days post-dose (300 mg dosing cohorts only). To determine AZD7442 concentrations in NLF, Nasosorption™ Fx-i (Mucosal Diagnostics, West Sussex, England) samples were collected 7 and 30 days post-dose. Preliminary interim results of serum and NLF antibody concentrations, and neutralizing antibody titers are reported to confirm preclinical information of AZD7442. The overall results of the Phase 1 study will be reported separately.

#### Nasal lining fluid pharmacokinetic assay

Qualified assays were used to measure the concentration of AZD7442 and urea in NLF eluent. NLF samples were collected by Nasosorption™ FX·i. To extract NLF, 300 µL of elution buffer (1.0 mg/mL BSA, 1% NP-40 in 1x PBS, pH = 7.4) was added to each synthetic absorptive matrix. Immediately upon elution, up to 60 μL of NLF eluant was used to measure the urea concentration with an enzymatic-colorimetric assay. Next, 180 μL of NLF eluant was subjected to immunocapture and enzymatic digestion treatments identical to the method described for serum sample analysis. The urea concentrations in NLF eluant and serum were used for normalization of AZD7442 concentrations in the NLF to correct for different extraction volumes.

#### SARS-CoV-2 plaque reduction neutralization test

Phase 1 clinical trial samples were evaluated using the SARS-CoV-2 plaque reduction neutralization test (PRNT) at Viroclinics Biosciences (Rotterdam, NLD). A standard number of SARS-CoV-2 infectious units were incubated with serial dilutions of sera. After a one-hour pre-incubation period of the virus, 100 μL of the mixture was added to the cells for 16–24 hours. After incubation, cells were formalin-fixed followed by incubation with a monoclonal antibody targeting the viral nucleocapsid protein, followed by a secondary anti-human IgG peroxidase conjugate and KPL TrueBlue™ substrate. Images of all wells were acquired by a CTL ImmunoSpot analyzer, equipped with software to quantitate the nucleocapsid-positive cells (virus signal). The 80% neutralization titer (PRNT_80_) was calculated as described previously (*47*).

#### Prediction of AZD7442 concentrations and neutralizing mAb titers in human serum

AZD7442 concentrations and neutralizing antibody titers in human serum were predicted beyond month 9 using a pharmacokinetic model that consisted of a central blood and peripheral distribution compartment with first-order absorption from the injection site into the central compartment, and first-order elimination from the central compartment.

### Bioanalytical assay to measure AZD7442 pharmacokinetics in NHP, human serum or human NLF

#### Serum pharmacokinetic assay

Separate validated assays with similar methodologies were used to measure the concentrations of AZD7442 in cynomolgus macaque and in human sera. A 20 μL sample was diluted and extracted with streptavidin magnetic beads coated with biotinylated RBD of SARS-CoV-2. The isolated analytes were subjected to denaturation, reduction, alkylation, and trypsin digestion. After digestion, the extract was fortified with stable isotope labelled peptide internal standard working solution. The final extract was analyzed via ultra-high performance liquid chromatography coupled with tandem mass spectrometry (UHPLC-MS/MS) with positive electrospray. A linear, 1/concentration² weighted, least-squares regression algorithm was used to quantify unknown samples.

### Statistical methods

Descriptive statistics were used to present data. GraphPad Prism software (versions 8.4.3 or higher) was used for data analysis and graph production.

## List of Supplementary Materials

### Materials and Methods

Figure S1. Hydrogen-deuterium exchange confirms that AZD8895 and AZD1061 bind distinct, non-overlapping epitopes on the SARS-CoV-2 spike protein RBD

Figure S2. In vitro assays confirm that AZD8895, AZD1061 and AZD7442 demonstrate reduced Fc effector function consistent with mAbs with TM substitutions

Figure S3. Human IgG concentrations in NHPs before and after SARS-CoV-2 challenge in prophylaxis or treatment settings

Figure S4. AZD7442 administration protects cynomolgus macaques against SARS-CoV-2 infection in prophylaxis settings

Figure S5. Serum neutralizing antibody titers in NHPs before or after SARS-CoV-2 challenge infection in prophylaxis or treatment settings

References: 47-57.

## Data Availability

Data underlying the findings described in this manuscript may be requested in accordance with AstraZenecas data sharing policy described at https://astrazenecagroup-dt.pharmacm.com/DT/Home.

## Acknowledgments

The authors would like to thank all investigators and researchers involved in this study, including the investigators on the Phase 1 clinical study Dr Muna Albayaty (AstraZeneca) and Dr Pablo ForteSoto (Parexel), and the other members of the AstraZeneca AZD7442 Study Group: Meina Liang, Ruipeng Mu, Jiaqi Yuan. The authors acknowledge the contribution of Todd Suscovich and SeromYx (Cambridge, MA, USA) where the in vitro Fc effector function assays were conducted, and Viroclinics Biosciences (Rotterdam, the Netherlands) where the Phase 1 clinical samples were evaluated.

This research was developed with funding from the Defense Advanced Research Projects Agency under HR011-18-3-001. The views, opinions, and/or findings expressed are those of the authors and should not be interpreted as representing the official views or policies of the Department of Defense or the U.S. government.

Medical writing support was provided by Lorna Forse, PhD, Matthew Young, DPhil, and Carl V Felton, PhD, and editorial support was provided by Nicola Mountford, all of Prime Global, Knutsford, UK, supported by AstraZeneca according to Good Publication Practice guidelines (Link).

## Author contributions

Conceptualization: Y-ML, PMM, NLK, RAG, MTE, YH, AIR

Methodology: Y-ML, PMM, RHA, RAG, EJK, NJB, RC, ASH, YH, AIR

Investigation: AA, SD, DJF, KRe, RR, KRo, KMT, OC, JB, LIC, JD, YH, AIK, YH, AIR

Visualization: Y-ML, PMM, RHA, MEA, KS, MTE Funding acquisition: NLK, MNP, MTE

Project administration: RC

Supervision: Y-ML, PMM, RHA, EJK, KS, NJB, AIR JEC, JMD, HB, NLK, MNP, MTE

Writing – original draft: Y-ML, MTE, YH, AIR Writing – review & editing: All

## Competing interests

YML, RHA, RAG, MEA, AA, SD, DJF, EJK, KRe, RR, KRo, KS, KMT, NJB, JB, LIC, YH, AIR, MNP and MTE are employees of, and hold or may hold stock in, AstraZeneca.

PMM, OC, JD, HB and NLK were employees of AstraZeneca at the time of this study.

NLK is currently an employee of and holds stock options of Eli Lilly.

HA, AIK, ASH and JMD have no conflicts to declare

JEC is a recipient of the 2019 Future Insight Prize from Merck KGaA, which supported part of this work with a grant; has served as a consultant for Luna Biologics, is a member of the Scientific Advisory Boards of CompuVax and Meissa Vaccines; is the founder of IDBiologics; and has received sponsored research agreements from Takeda Vaccines, IDBiologics and AstraZeneca.

Opinions, discussions, conclusions, interpretations, and recommendations are those of the authors and are not necessarily endorsed by the U.S. Army. The mention of trade names or commercial products does not constitute endorsement or recommendation for use by the Department of the Army or the Department of Defense.

## Data and materials availability

Data underlying the findings described in this manuscript may be requested in accordance with AstraZeneca’s data sharing policy described at https://astrazenecagroup-dt.pharmacm.com/DT/Home.

