## Supplemental material for "AZD7442 demonstrates prophylactic and therapeutic efficacy in non-human primates and extended half-life in humans"

26 August 2021

### Supplementary Materials

#### Supplementary methods

##### Hydrogen-deuterium exchange mass spectrometry

125 µl SPIKE stock (4.3 µM trimer) was added to 10 µl IgG stock (~10 mg/ml) to give a molar ratio of 1:1.2 (SPIKE: IgG) of either AZD1061 or AZD8895 and equilibrated for 30 min at room temperature. LEAP hydrogen-deuterium exchange (HDX) method: 7.6 µL of protein sample (either 4.3 µM SPIKE stock [control], or 1:1.2 SPIKE and IgG sample) was added to 52.4 µL label buffer (10 mM potassium phosphate pD 6.2) and incubated for either 1, 10 or 100 min at 20°C. After incubation, 50 µL of this sample was added to 50 µl quench solution (100 mM potassium phosphate, 200 mM TCEP, 2M Gdn-HCl, pH 2.3). 95 µL of the quenched sample was injected onto the HDX manager (50 µL loop volume). LC-MS method: Online digestion was performed at 20°C via an online pepsin column (Waters, enzymate) at 100 µL/min for 4 mins (~4800 psi). After trapping peptides were separated via a C18 analytical column at 40 µL/min using a 5-35% linear MeCN gradient over 6 minutes. The MS analyser was a Thermo orbitrap fusion instrument operating in orbitrap detection mode at a resolution of 120k. Data were processed using biopharma finder v3.0.62.11, with a S/N threshold of 256 and a mass error threshold of 15 ppm. HDX data were processed using HDExaminer version 2, excluding the N-terminal residue of each peptide due to back exchange. Structural heat maps were generated using PAVED (*49*).

This document has not been peer reviewed

NOTE: This preprint reports new research that has not been certified by peer review and should not be used to guide clinical practice.

#### Protein expression and purification

Codon-optimized genes were cloned into mammalian expression vectors. Monoclonal antibodies (mAbs) were IgG1 constructs with wild-type (WT) fragment crystallizable (Fc), or Fc variants containing TM and YTE substitutions. The YTE substitution (M252Y/S254T/T256E) in the Fc region of immunoglobulin G (IgG) has been shown to increase affinity for the human neonatal Fc receptor (huFcRn) at lower endosomal pH (*40*), promoting recirculation and thereby extending the elimination half-life ($t_{1/2}$) of mAbs in humans by 85–117 days (*20, 39, 50-52*). The TM substitution (L234F/L235E/P331S) in the Fc region of IgG has been shown to eliminate antibody-dependent cellular cytotoxicity and complement-dependent cytotoxicity in clinical settings (*22, 53-55*).

SARS-CoV-2 spike variant constructs were designed using the sequence of the Wuhan-Hu-1 (GenBank: MN908947.3) isolate as a reference. SARS-CoV-2 receptor binding domain (RBD) (residues 334–526) constructs were cloned with N-terminal CD33 leader sequences and C-terminal GSSG linkers, AviTags, GSSG linkers, and 8x HisTags. SARS-CoV-2 NTD (residues 16–305) was cloned with an N-terminal CD33 leader sequence and C-terminal GSSG linker and 8x HisTag. SARS-CoV-2 trimers were designed as previously described (*48, 56*). The human *ACE2* gene (residues 19–615) was cloned with an N-terminal CD33 leader sequence and either a C-terminal GSSG linker, AviTag, GSSG linker and 8x HisTag, or a C-terminal GSSG linker and 8x HisTag. Proteins were expressed in FreeStyle 293X or 293F cells transfected with 293fectin (Thermo Fisher Scientific, Waltham, MA, USA). mAbs were isolated from supernatants

This document has not been peer reviewed

NOTE: This preprint reports new research that has not been certified by peer review and should not be used to guide clinical practice.

by affinity chromatography using MabSelect or Protein A or Protein G columns (GE Healthcare), eluted with 0.1 M glycine at low pH, then dialyzed into PBS. Other proteins were isolated by affinity chromatography using a HisTrap column followed by size exclusion column chromatography (GE Healthcare). Purified proteins were pooled and analyzed by gel electrophoresis, and further evaluated by size-exclusion chromatography coupled to multi-angle light scattering for some proteins. Protein biotinylation was performed enzymatically with BirA biotin protein ligase (Avidity, Aurora, CO, USA) and confirmed by Western blot or HABA reagent (ThermoFisher Scientific).

This document has not been peer reviewed

#### Binding assays

##### *Antibody binding assay*

Binding assays were performed by enzyme-linked immunosorbent assay (ELISA) using Nunc MaxiSorp 384-well plates coated with 20 µL/well of antigen. Antigens were diluted in PBS (pH 7.2) and coating concentrations were 5 µg/mL for RBD variants, NTD, and hemagglutinin and 3 µg/mL for spike trimers. Coated plates were incubated overnight at 4°C. Plates were washed 6 times with PBST and then wells were blocked with 100 µL of casein buffer for 1 h at room temperature (RT). Blocking buffer was removed and 20 µL of serially diluted antibody (diluted in casein in a separate plate at a starting concentration of 10 µg/mL) was added to wells in duplicate and incubated 1 h at RT with shaking. Plates were washed 6 times with PBST and then 20 µL of anti-human-Fc-HRP secondary (diluted 20k in PBST in separate tube) was added to each well and incubated for 30 min at RT with shaking before washing again with PBST. Plates were

NOTE: This preprint reports new research that has not been certified by peer review and should not be used to guide clinical practice.

developed with TMB added at 20 μL/well and incubated 5 mins at RT before acid quenching with 10 μL/well of 2N $H_2SO_4$. Wells were measured for absorbance at 450 nm using an EnVision 2105 Multimode Plate Reader (Perkin Elmer, Boston, MA, USA). Absorbance values were graphed with GraphPad Prism software (version 8.4.2) and data were fit to a one-site binding curve. Sample additions for blocking, secondary, and development steps were all performed via Multidrop (Thermo Fisher Scientific). Assays were performed twice in duplicate.

***Antibody binding kinetics***

Kinetic rates of AZD1061 and AZD8895 binding to SARS-CoV-2 spike trimer and spike RBD were evaluated by surface plasmon resonance technology (Biacore T200). Affinity to SARS-Cov-2 spike trimer was determined by first immobilizing murine anti-his Ab to a net response of ~5,000 RUs onto a CM5 chip, then capturing SARS-CoV-2 trimer and measuring binding of titrated IgG and Fab of AZD8895, AZD1061, or AZD7442 (on for 100 s and off for 500 s at 50 μL/min). Affinity to SARS-Cov-2 spike RBD was determined by immobilizing Protein A to a net response of ~3,000 RUs onto a CM5 chip, then capturing IgGs and measuring binding to titrated SARS-CoV-2 spike RBD-Avi-His (on for 100 s and off for 1,000 s at 50 μL/min for AZD1061; on for 50 s and off for 200 s at 50 μL/min for AZD8895). Measuring ACE2 affinity to SARS-Cov-2 spike trimer was determined by immobilizing strep-tactin XT to a net response of ~2,400 RUs onto a CM5 chip, then capturing ~250 RUs SARS-CoV-2 S protein trimer and measuring binding to titrated ACE2-Avi-His (on for 100 s and off for 200 s at 50 μL/min).

NOTE: This preprint reports new research that has not been certified by peer review and should not be used to guide clinical practice.

This document has not been peer reviewed

Collected data were fit to a one-site binding equation using Biacore software to obtain binding kinetics measurements.

##### *SARS-CoV-2 receptor binding inhibition assay*

A modified ELISA was conducted to measure mAb inhibition potencies for blocking RBD binding to ACE2. Ninety-six-well plates were coated overnight at 4°C with 100 µL/well of Penta-His antibody diluted in PBS (pH 7.0) to 1 µg/mL final concentration. Plates were washed with PBST, and then wells were blocked with 250 µL casein for 1 h at RT. Blocking buffer was removed from plates and ACE2-His was captured by addition of 100 µL/well ACE2-His diluted in casein to 1 µg/mL. Plates were incubated for 2 h at RT. During this time, mAbs were serially diluted in casein, starting at a concentration of 20 µg/mL. The mAb dilutions were then mixed with an equal volume of RBD-Fc (diluted to 0.1 µg/mL) and the mAb-RBD mixtures incubated for 1 h at RT. Plates were washed with PBST to remove unbound ACE2-His before addition of 100 µL/well of the mAb-RBD mixtures. After incubation for 1.5 h at RT, unbound protein was removed by washing with PBST. Bound RBD-Fc was detected by the addition of 100 µL/well HRP-conjugated goat anti-human IgG Fc antibody (diluted 1:55,000 in PBST). Plates were incubated for 1 hr at RT, washed again with PBST and then developed with the addition of 100 µL/well TMB. Plates developed for 5 min at RT before acid quenching with 50 µL/well 2N $H_2SO_4$. Wells were measured for absorbance at 450 nm using an EnVision 2105 Multimode Plate Reader (Perkin Elmer). Absorbance values were fit to nonlinear regression curve fit (four parameters) using GraphPad Prism software (version 8.4.3).

NOTE: This preprint reports new research that has not been certified by peer review and should not be used to guide clinical practice.

This document has not been peer reviewed

Sample additions for blocking, secondary, and development steps were all performed via Multidrop (Thermo Fisher Scientific). The assay was performed twice in duplicate.

##### *mAb binding to human FcRn, FcγR, and complement C1q*

Binding kinetics of mAbs to human FcRn, FcγRs, and complement C1q were assessed by SPR using a Biacore T200. FcRn binding to the mAbs was measured at pH 6.0 and 7.4 and at 750 nM (2000 s at 5 μL/min). The assay was performed in duplicate. mAb binding $K_D$ values were determined at pH 6.0 by fitting data to a 1:1 binding isotherm equation with Biacore evaluation software (Cytiva, Marlborough, MA, USA). pH dependency was calculated as (1 – (RU pH 7.4 / RU pH 6.0)) × 100.

Human FcγRs was immobilized to a net response of 3,000-8,000 RU onto a CM5 sensor chip; C1q protein was immobilized to a net response of 5,000 RU onto a CM4 chip. AZD8895, AZD8895-TM, AZD1061, and AZD1061-TM binding was measured at physiological mAb concentrations observed in human serum at ~50, 150, or 450 μg/mL (333, 1,000, or 3,000 nM; 2,000 s at 10 μL/min) with final response measurements at each sample dilution used to determine binding as a function of concentrations. Assays were performed in duplicate, and data was fit to a one site binding equation in GraphPad Prism.

This document has not been peer reviewed

NOTE: This preprint reports new research that has not been certified by peer review and should not be used to guide clinical practice.

#### Virus neutralization assays

##### *SARS-CoV-2 microneutralization assays*

SARS-CoV-2 reference strains (USA-WA1/2020 or AUS/VIC01/2020) and VOC (B.1.1.7, B.1.351, P.1, and B.1.617.2) were obtained from BEI Resources (Manassas, VA, USA) or isolated from nasopharyngeal swabs obtained and expanded in Vero/E6 cells. All viruses were shown to be identical to the described genotype of circulating viruses by whole genome sequencing. Microneutralization assays at Public Health England (PHE; Porton Down, UK), United States Army Medical Research Institute of Infectious Diseases (USAMRIID; Frederick, MD, USA), and Integrated Research Facilitity, National Institute of Allergy and Infectious Diseases (IRF/NIAID; Frederick, MD, USA) were used to measure mAb potency by incubating infectious virus with serially diluted mAbs (AZD8895, AZD1061, and AZD7742). Virus-susceptible monolayers (Vero/E6 cells) were exposed to the mAb/virus mixture. Following fixation, cells were immunostained with antibodies specific for SARS-CoV-2 nucleocapsid or spike RBD (Sino Biological, Beijing, China). Infected cells were detected using either an HRP-labeled secondary antibody with KPL TrueBlue™ substrate for visualization, or an AlexaFluor 488-labeled secondary antibody with high content fluorescence imaging for visualization. $IC_{50}$ values were determined from non-linear regression analysis. Relative reduction in neutralization potencies (fold-change $IC_{50}$) of mAbs against VOC was determined relative to reference strains tested in parallel.

NOTE: This preprint reports new research that has not been certified by peer review and should not be used to guide clinical practice.

This document has not been peer reviewed

##### *SARS-CoV-2 plaque reduction neutralization test*

Serum samples from cynomolgus macaques were evaluated using the SARS-CoV-2 plaque reduction neutralization test (PRNT) at Viroclinics Biosciences (Rotterdam, the Netherlands) and BIOQUAL, Inc. (Rockville, MD, USA), respectively. A standard number of SARS-CoV-2 infectious units were incubated with serial dilutions of sera. After a one-hour pre-incubation period of the serum mixtures, 100 µL of the mixture was added to the cells for 16–24 hours. After incubation, cells were formalin-fixed followed by incubation with a mAb targeting the viral nucleocapsid protein, followed by a secondary anti-human IgG peroxidase conjugate and KPL TrueBlue™ substrate. Images of all wells were acquired by a CTL ImmunoSpot analyzer, equipped with software to quantitate the nucleocapsid-positive cells (virus signal). The median neutralization titer ($PRNT_{50}$) was calculated as described previously (*47*).

##### ***In vitro*** **Fc effector function studies**

This document has not been peer reviewed

##### *Chemical conjugation of SARS-CoV-2 spike trimer to fluorescent polystyrene beads*

Fluorescent polystyrene beads (Thermo Fisher Scientific) were washed in sterile cell culture-grade water and centrifuged at 15,000 × *g* to pellet. The beads were resuspended in 100 mM sodium phosphate buffer pH 6.2. Sulfo-NHS reconstituted at 50 mg/mL in sterile cell culture-grade water and 1-ethyl-3- [3-dimethylaminopropyl] carbodiimide reconstituted at 50 mg/mL in 100 mM sodium phosphate buffer pH 6.2

NOTE: This preprint reports new research that has not been certified by peer review and should not be used to guide clinical practice.

(activation buffer) were added to beads and vortexed thoroughly to mix. The beads were incubated at RT on a rotator. After the incubation, the beads were centrifuged at 15,000 × *g* and the supernatant was discarded. The bead pellet was resuspended in 50 mM MES buffer pH 5.0 (coupling buffer) and vortexed thoroughly to mix. The beads were centrifuged at 15,000 × *g*, supernatant discarded, and washed in coupling buffer for a total of three washes. Beads were pelleted after final wash and resuspended in 50 mM MES buffer pH 5.0. Approximately 25 µg of SARS-CoV-2 spike trimer was added to the beads and incubated for 2 h at RT on a rotator. Beads were pelleted at 15,000 × *g* and resuspended in PBS-Tween (blocking/storage buffer) and incubated for 30 mins at RT on a rotator. Beads were pelleted at 15,000 × *g* and washed three times in PBS-Tween (wash buffer). After the final wash, beads were centrifuged at 15,000 × *g* and resuspended in PBS. The coupled fluorescent beads were stored at 4$^{o}$C in the dark until use.

This document has not been peer reviewed

***Antibody-dependent cellular phagocytosis with THP-1 cells***

Antibody-dependent cellular phagocytosis (ADCP) assesses the ability of antibodies to induce phagocytosis of antigen-functionalized fluorescent beads by monocytes via Fc receptors. Fluorescent, streptavidin-conjugated polystyrene beads were coated with biotinylated SARS-CoV-2 spike trimer. Diluted antibody (in PBS) was added, and unbound antibodies were washed away. The antibody:bead complexes were added to undifferentiated THP-1 cells (ATCC, Manassas, VA, USA), a monocytic cell line, and phagocytosis was allowed to proceed overnight. The cells were then washed and fixed,

NOTE: This preprint reports new research that has not been certified by peer review and should not be used to guide clinical practice.

and the extent of phagocytosis was measured by flow cytometry. Data are reported as a phagocytic score, which considers the proportion of effector cells that phagocytosed and the degree of phagocytosis. The mAbs were tested for ADCP activity at a range of 5 µg/mL to 2.3 ng/mL.

***ADCP with neutrophils***

ADCP assesses the ability of antibodies to induce the phagocytosis of antigen-coated targets by primary neutrophils. Fluorescent, streptavidin-conjugated polystyrene beads were coupled to biotinylated SARS-CoV-2 spike trimer. Diluted antibody (in PBS) was added, and unbound antibodies were washed away. The antibody:bead complexes were added to primary neutrophils isolated from healthy blood donors using negative selection (StemCell EasySep Direct Human Neutrophil Isolation Kit), and phagocytosis was allowed to proceed for 1 h. The cells were then washed and fixed, and the extent of phagocytosis was measured by flow cytometry. Data are reported as a phagocytic score, which considers the proportion of effector cells that phagocytosed and the degree of phagocytosis. Each sample was run in biological duplicate using neutrophils isolated from two distinct donors. The mAbs were tested for ADNP activity at a range of 67 µg/mL to 30.6 ng/mL.

This document has not been peer reviewed

***Antibody-dependent cellular cytotoxicity***

Antibody-dependent cellular cytotoxicity (ADCC) tests the ability of antigen-specific antibodies to recruit NK cell lytic activity. Target cells were biotinylated (EZ-Link Sulfo-

NOTE: This preprint reports new research that has not been certified by peer review and should not be used to guide clinical practice.

NHS-LC-Biotin) and stained with one of two dyes (half with CellTrace Violet and half with CellTrace Far Red). The stained cells were then pulsed with either streptavidin-conjugated stabilized SARS-CoV-2 spike trimer or left unpulsed. Diluted antibody (in PBS) was added to a 1:1 mixture of the target cells stained with each dye. NK cells purified from healthy blood donor leukopaks using commercially available negative selection kits (EasySep Human NK Cell Isolation Kit, StemCell, Cambridge, MA, USA) were added and incubated for 4 h. Following this incubation, the cells were stained with a viability dye (Zombie Green Fixable Viability Kit, BioLegend, San Diego, CA, USA). Lysis was measured by flow cytometry and reported as the percent of dead (viability dye positive) antigen-coated cells. The mAbs were tested for ADCC activity at a range of 25 µg/mL to 1.5 ng/mL.

This document has not been peer reviewed

***Antibody-dependent complement deposition***

Antibody-dependent complement deposition (ADCD) assesses the recruitment of complement component C3b on the surface of antigen-coupled beads. Neutravidin-conjugated polystyrene beads were coated with biotinylated SARS-CoV-2 spike trimer. Diluted antibody (in PBS) was added, and unbound antibodies were washed away. Commercially available guinea pig complement was then added as a source of complement. Following a brief incubation, the complement was washed away, and a fluorescein isothiocyanate (FITC)-conjugated mAb specific for guinea pig C3 was added. Complement deposition was measured using flow cytometry and was reported

NOTE: This preprint reports new research that has not been certified by peer review and should not be used to guide clinical practice.

as the median fluorescent intensity of FITC. The mAbs were tested for ADCD activity at a range of 100 μg/mL to 4646 ng/mL.

##### *Antibody-dependent enhancement of infection*

The antibody-dependent enhancement of infection (ADEI) assay measures the capacity of antibodies to facilitate the infection of FCGR-expressing cells. Antibodies were diluted in cell culture medium, and SARS-CoV-2 spike protein-pseudotyped lentiviruses encoding firefly luciferase were added. The antibodies and viruses were incubated for 1 h, after which Raji cells (a B-cell line that naturally expresses FCGR2; ATCC, Manassas, VA, USA) were added. The cultures were incubated for 24 h, after which the culture medium was removed, and luciferase assay reagent was added. Following a 2-min incubation to allow for cell lysis, the supernatant was transferred to a black 96-well plate, and luciferase activity was measured using a luminometer. Data are reported as the relative infection rate compared to infection in the absence of antibody. The mAbs were tested for ADEI activity at a range of 1 μg/mL to 0.125 μg/mL.

This document has not been peer reviewed

##### *Antibody-dependent NK cell activation*

Antibody-dependent natural killer cell activation (ADNKA) assesses antigen-specific antibody-mediated natural killer (NK) cell activation against protein-coated plates. Stabilized SARS-CoV-2 spike trimer was used to coat ELISA plates, which were then washed and blocked. Diluted antibody (in PBS) was added to the antigen-coated plates, and unbound antibodies were washed away. NK cells, purified from healthy blood donor

NOTE: This preprint reports new research that has not been certified by peer review and should not be used to guide clinical practice.

leukopaks using commercially available negative selection kits (StemCell EasySep Human NK Cell Isolation Kit) were added, and the levels of activation marker (CD107a) and intracellular cytokines (IFN-γ and MIP-1β) were measured after 5 h using flow cytometry. Data are reported as the percentage of cells positive for each of the activation markers (CD107a, IFN-γ and MIP-1β). Each sample was tested with at least two different NK cell donors, with all samples tested with each donor. The mAbs were tested for ADNKA activity at a range of 20 μg/mL to 9.1 ng/mL.

This document has not been peer reviewed

NOTE: This preprint reports new research that has not been certified by peer review and should not be used to guide clinical practice.

26 August 2021

### Supplementary results

#### Figure S1. Hydrogen-deuterium exchange confirms that AZD8895 and AZD1061 bind distinct, non-overlapping sites on the SARS-CoV-2 spike protein RBD

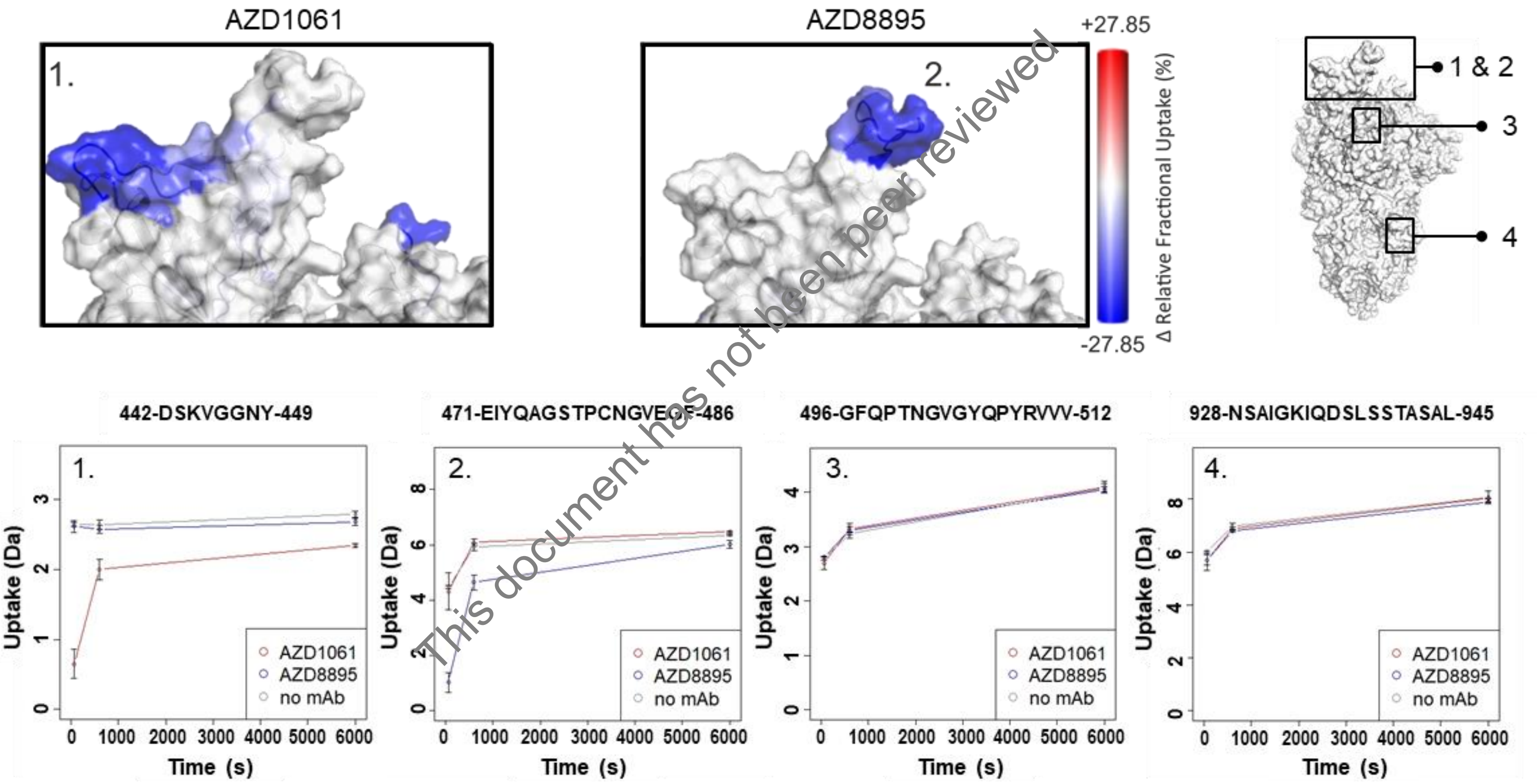


This document has not been peer reviewed

NOTE: This preprint reports new research that has not been certified by peer review and should not be used to guide clinical practice.

Structural heat map of HDX data reveals relative protection from HD exchange in distinct, complementary regions of the RBD following binding of AZD1061 (box 1, top left) or AZD8895 (box 2, top right). Below panels: example uptake plots of peptides covering these regions (numbered 1 and 2 for the sites of AZD1061 and AZD8895, respectively) as well as two other peptides distal to these sites showing similar deuterium uptake behavior (peptides 3 and 4, as highlighted on the SPIKE protein structure in the top left) are shown for SPIKE alone (cyan), SPIKE + AZD1061 (black) and SPIKE + AZD8895 (yellow). Error bars show standard deviation (SD), n=3. PDB 6Z97 (*57*).

mAb, monoclonal antibody; RBD, receptor-binding domain; SARS-CoV-2, severe acute respiratory syndrome coronavirus 2; SD, standard deviation.

This document has not been peer reviewed

NOTE: This preprint reports new research that has not been certified by peer review and should not be used to guide clinical practice.

26 August 2021

**Figure S2. *In vitro* assays confirm that AZD8895, AZD1061 and AZD7442 demonstrate reduced Fc effector function consistent with mAbs with TM substitutions**

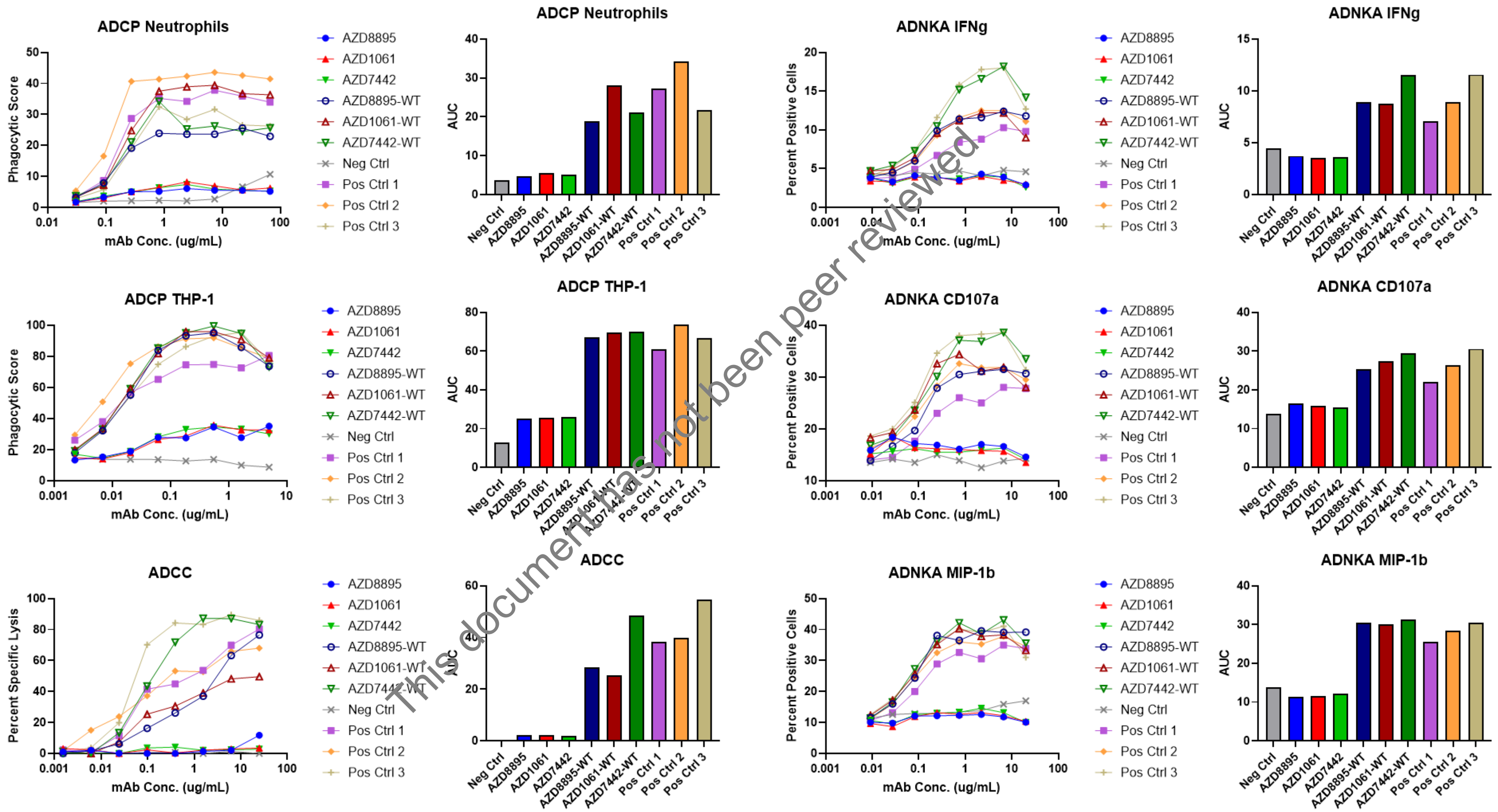


This document has not been peer reviewed

NOTE: This preprint reports new research that has not been certified by peer review and should not be used to guide clinical practice.

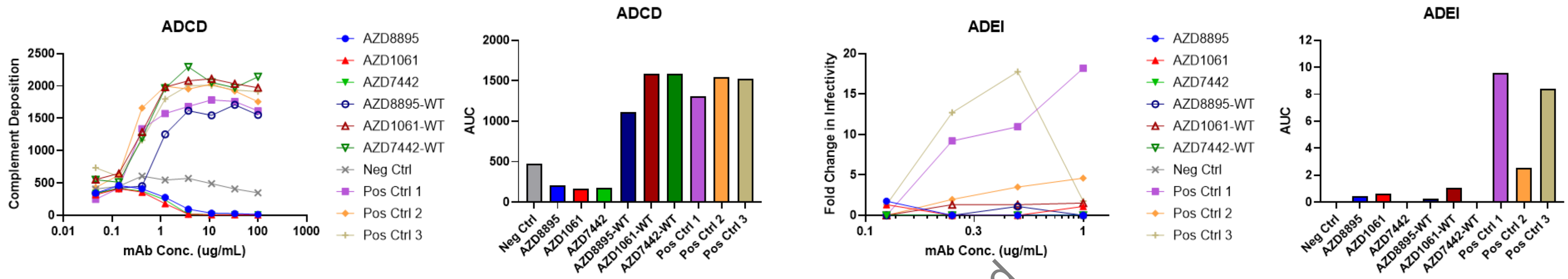


*In vitro* ADCP, ADCC, ADNKA, ADCD and ADEI assays were conducted with mAbs diluted across a range of concentrations. For ADCP assays, effector cells were measured for the uptake of spike-bearing fluorescent beads in the absence or presence of mAbs by flow cytometry. For ADCC assay, spike-expressing target cells were measured for neutrophil-mediated killing in the absence or presence of mAbs by flow cytometry. For ADNKA assay, primary NK cells were exposed to spike-coated plates in the absence or presence of mAbs and flow cytometry analyses conducted to measure expression of activation markers and cytokines. For ADCD assay, flow cytometry analyses were conducted to measure the deposition of complement proteins onto spike-coated beads in the absence or presence of mAbs. For ADEI assay, Raji cells were incubated with a spike-expressing pseudovirus that encoded a luciferase reporter. Incubations were performed in the absence or presence of mAbs with virus uptake measured by luciferase expression. Graphs show mean response subtracting background from 2 or more independent experiments. Bar charts show AUC analysis for each graph.

This document has not been peer reviewed

NOTE: This preprint reports new research that has not been certified by peer review and should not be used to guide clinical practice.

ADCC, antibody-dependent cell cytotoxicity; ADCD, antibody-dependent complement deposition; ADCP, antibody-dependent cellular phagocytosis; ADEI, antibody-dependent enhancement of infection, ADNKA, antibody-dependent NK activation; AUC, area under the curve, CD, cluster of differentiation; Ctrl, control; Fc, fragment crystallizable; IFNg, interferon gamma; mAb, monoclonal antibody; MIP-1b, macrophage inflammatory protein also known as CCL4; Neg, negative; Pos, positive; THP-1, human monocytic cell line

This document has not been peer reviewed

NOTE: This preprint reports new research that has not been certified by peer review and should not be used to guide clinical practice.

26 August 2021

**Figure S3. Human IgG concentrations in sera of NHPs before and after SARS-CoV-2 challenge with antibodies administered in prophylaxis or treatment settings**

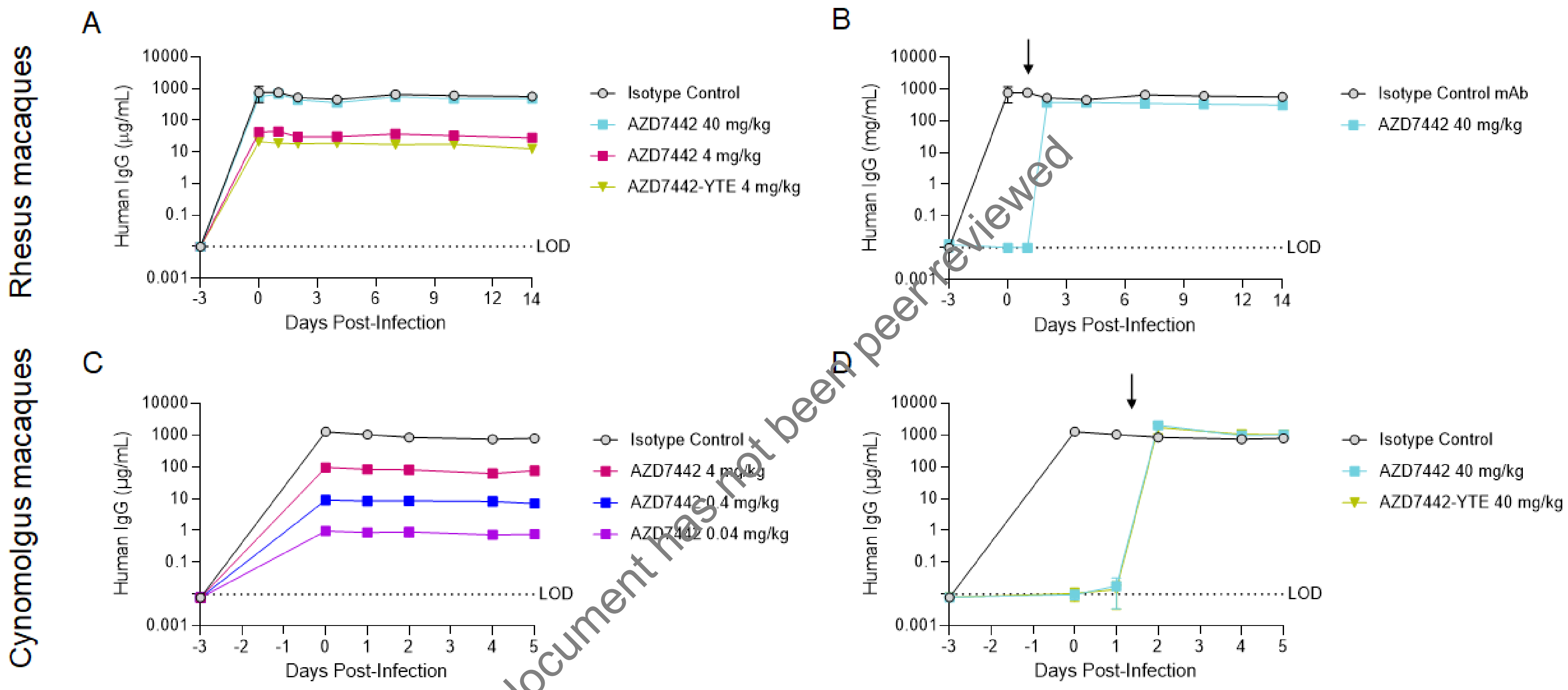


This document has not been peer reviewed

NOTE: This preprint reports new research that has not been certified by peer review and should not be used to guide clinical practice.

Rhesus macaques in prophylaxis Groups 1, 2, 3, and 4 (n=3 for Groups 1 and 2; n=4 for Groups 3 and 4) received an IV infusion of mAb 3 days prior to challenge. Rhesus macaques in treatment Group 5 (n=4) received an IV infusion of mAb 1 day after challenge. Rhesus macaques were challenged with $10^5$ PFU of SARS-CoV-2, split between IT and IN delivery on Day 0. Serum was collected on Days -3, 0, 1, 2, 4, 7, 10 and 14. Cynomolgus macaques in prophylaxis Groups 1, 2, 3, and 4 (all groups n=3), received an IV infusion of mAb three days prior to challenge. Cynomolgus macaques in treatment Groups 5 and 6 (n=3) received an IV infusion of mAb one day after challenge. Cynomolgus macaques were challenged with $10^5$ $TCID_{50}$ of SARS-CoV-2, split between IT and IN delivery on Day 0. Serum was collected on Days -3, 0, 1, 2, 4, and 5.

Data are geometric mean ± SD human serum IgG concentrations following IV administration of isotype control mAb, AZD7442 or AZD7442-YTE in (A, B) rhesus macaques or (C, D) cynomolgus macaques, administered either (A, C) prophylactically or (B, D) as treatment. One animal in the rhesus macaque control group (Group 1) missed being dosed with the isotype control mAb, and as expected, showed no detectable levels of circulating human mAb, and was excluded from this analysis of human IgG concentration in serum.

D, day relative to SARS-CoV-2 challenge (Day 0); IgG, immunoglobulin G; IN, intranasal; IT, intratracheal; IV, intravenous; LOD, limit of detection; mAb, monoclonal antibody; NHP, non-human primate; SARS-CoV-2, severe acute

This document has not been peer reviewed

NOTE: This preprint reports new research that has not been certified by peer review and should not be used to guide clinical practice.

respiratory syndrome coronavirus 2; PFU, plaque-forming unit; SD, standard deviation; YTE, Immunoglobulin constant heavy chain substitution to extend mAb half-life.

This document has not been peer reviewed

NOTE: This preprint reports new research that has not been certified by peer review and should not be used to guide clinical practice.

26 August 2021

**Figure S4. AZD7442 administration protects cynomolgus macaques against SARS-CoV-2 infection in prophylaxis settings**

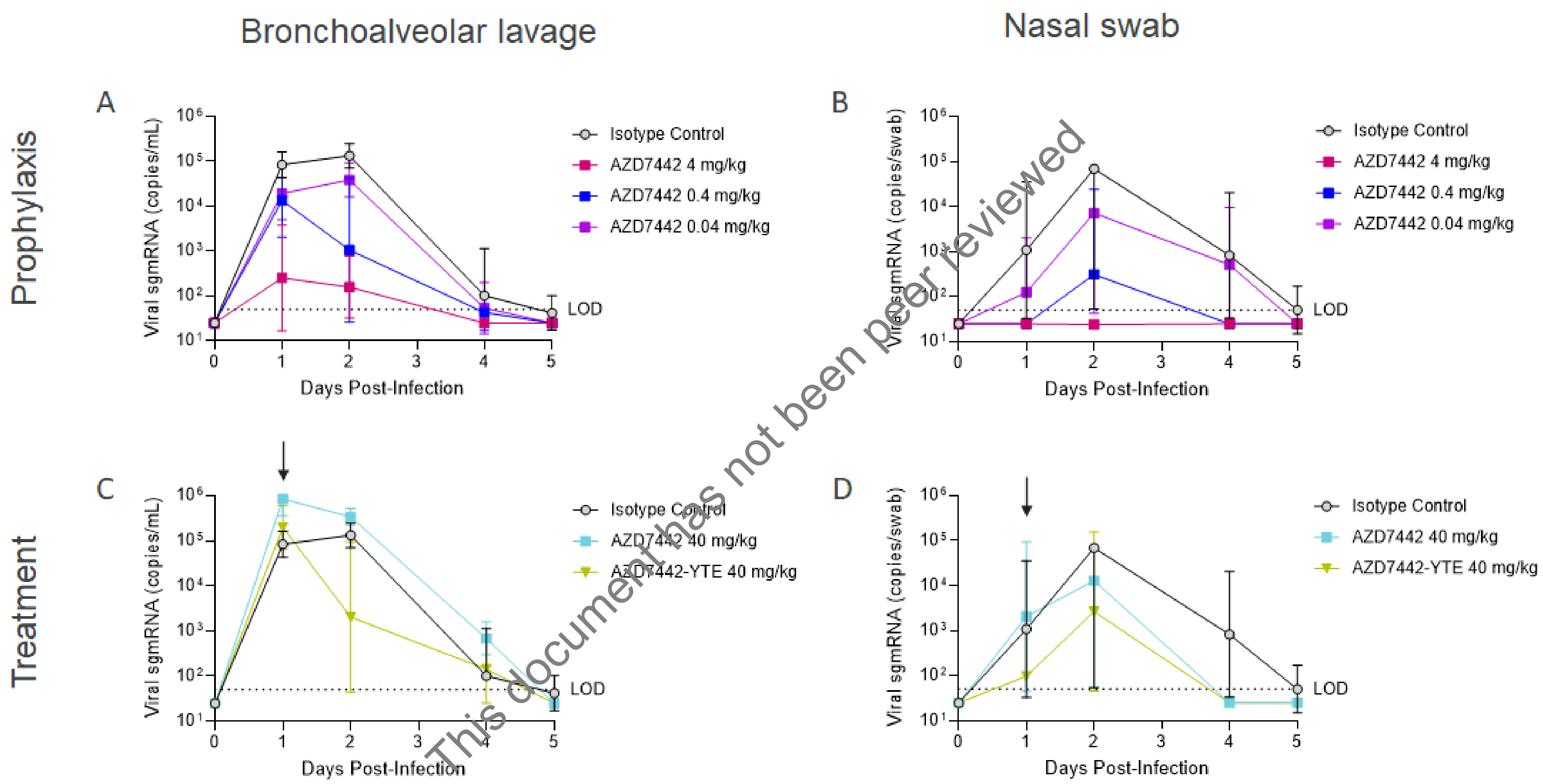


This document has not been peer reviewed

NOTE: This preprint reports new research that has not been certified by peer review and should not be used to guide clinical practice.

Cynomolgus macaques in prophylaxis Groups 1, 2, 3, and 4 (all groups n=3) received IV infusions of mAb three days prior to challenge. Cynomolgus macaques in treatment Groups 5 and 6 (n=3) received an IV infusion of mAb 1 day after challenge. Cynomolgus macaques were challenged with $10^5$ TCID50 of SARS-CoV-2, split between IT and IN delivery on Day 0. BAL and nasal swab samples were collected at Days 0, 1, 2, 4, 7, 10, and 14.

Data are geometric mean ± SD viral burden following IV administration of isotype control mAb, AZD7442 or AZD7442-YTE in a **(A, B)** prophylaxis or **(C, D)** treatment setting, with samples taken from **(A, C)** BAL or **(B, D)** nasal swabs

BAL, bronchoalveolar lavage; D, day; IN, intranasal; IT, intratracheal; IV, intravenous; mAb, monoclonal antibody; sgmRNA, subgenomic messenger RNA; SARS-CoV-2, severe acute respiratory syndrome coronavirus 2; SD, standard deviation.

This document has not been peer reviewed

NOTE: This preprint reports new research that has not been certified by peer review and should not be used to guide clinical practice.

26 August 2021

**Figure S5. Serum neutralizing antibody titers in NHPs before or after SARS-CoV-2 challenge infection in prophylaxis or treatment settings**

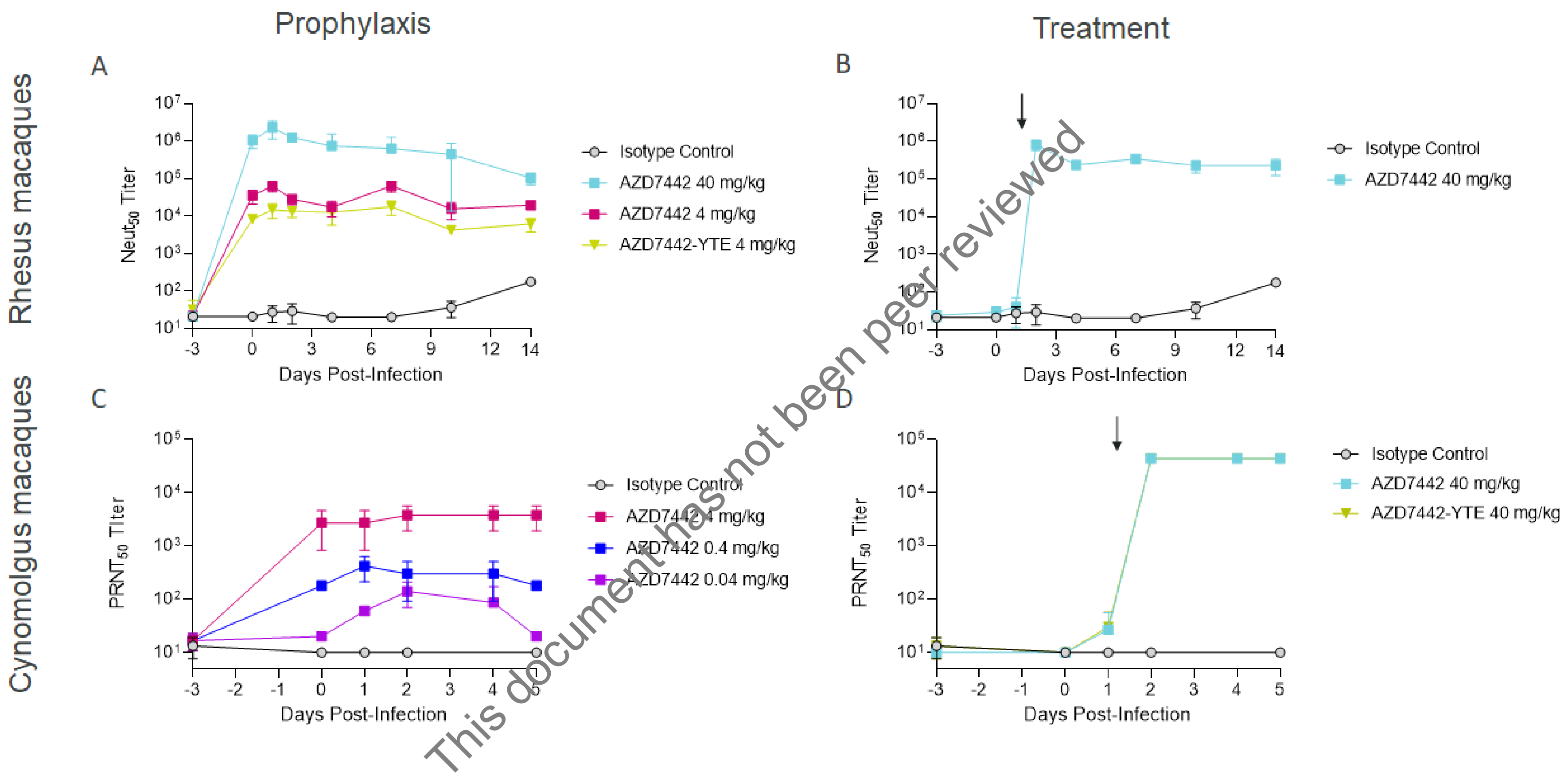


This document has not been peer reviewed

NOTE: This preprint reports new research that has not been certified by peer review and should not be used to guide clinical practice.

Rhesus macaques in prophylaxis Groups 1, 2, 3, and 4 (n=3 for Groups 1 and 2; n=4 for Groups 3 and 4) received an IV infusion of mAb 3 days prior to challenge. Rhesus macaques in treatment Group 5 (n=4) received an IV infusion of mAb 1 day after challenge. Rhesus macaques were challenged with $10^5$ PFU of SARS-CoV-2, split between IT and IN delivery on Day 0. Serum was collected on Days -3, 0, 1, 2, 4, 7, 10 and 14. Cynomolgus macaques in prophylaxis Groups 1, 2, 3, and 4 (all groups n=3), received an IV infusion of mAb 3 days prior to challenge. Cynomolgus macaques in treatment Group 5 (n=3) received an IV infusion of mAb 1 day after challenge. Cynomolgus macaques were challenged with $10^5$ $TCID_{50}$ of SARS-CoV-2, split between IT and IN delivery on Day 0. Serum was collected on Days -3, 0, 1, 2, 4, and 5.

Data shown is geometric mean ± SD serum neutralizing antibody titers following IV administration of isotype control mAb, AZD7442 or AZD7442-YTE in **(A, B)** rhesus macaques or **(C, D)** cynomolgus macaques, administered either **(A, C)** prophylactically 3 days prior to SARS CoV-2 challenge, or **(B, D)** as treatment 1 day after SARS CoV-2 infection. Neutralizing antibody titers were measured using a SARS-CoV-2 spike pseudovirus neutralization assay **(A, B)** or a SARS-CoV-2 plaque reduction neutralization test **(C, D)**.

IV, intravenous; mAb, monoclonal antibody; NHP, non-human primate; PFU, plaque-forming unit; SARS-CoV-2, severe acute respiratory syndrome coronavirus 2; SD, standard deviation.

This document has not been peer reviewed

NOTE: This preprint reports new research that has not been certified by peer review and should not be used to guide clinical practice.
